# Epidemiology of *Plasmodium malariae* and *Plasmodium ovale* spp. in a highly malaria-endemic country: a longitudinal cohort study in Kinshasa Province, Democratic Republic of Congo

**DOI:** 10.1101/2023.04.20.23288826

**Authors:** Rachel Sendor, Kristin Banek, Melchior Mwandagalirwa Kashamuka, Nono Mvuama, Joseph A. Bala, Marthe Nkalani, Georges Kihuma, Joseph Atibu, Kyaw L. Thwai, W. Matthew Svec, Varun Goel, Tommy Nseka, Jessica T. Lin, Jeffrey A. Bailey, Michael Emch, Margaret Carrel, Jonathan J. Juliano, Antoinette Tshefu, Jonathan B. Parr

**Author notes:** Corresponding author: (RS). AT and JBP are joint senior authors.

## Abstract

**Background:** Increasing reports suggest that non-falciparum species are an underappreciated cause of malaria in sub-Saharan Africa, but their epidemiology is not well-defined. This is particularly true in regions of high *P. falciparum* endemicity such as the Democratic Republic of Congo (DRC), where 12% of the world’s malaria cases and 13% of deaths occur.

**Methods and Findings:** The cumulative incidence and prevalence of P. malariae and P. ovale spp. infection detected by real-time PCR were estimated among children and adults within a longitudinal study conducted in seven rural, peri-urban, and urban sites from 2015-2017 in Kinshasa Province, DRC. Participants were sampled at biannual household survey visits (asymptomatic) and during routine health facility visits (symptomatic). Participant-level characteristics associated with non-falciparum infections were estimated for single- and mixed-species infections. Among 9,089 samples collected from 1,565 participants over a 3-year period, the incidence of P. malariae and P. ovale spp. infection was 11% (95% CI: 9%-12%) and 7% (95% CI: 5%-8%) by one year, respectively, compared to a 67% (95% CI: 64%-70%) one-year cumulative incidence of P. falciparum infection. Incidence continued to rise in the second year of follow-up, reaching 26% and 15% in school-age children (5-14yo) for P. malariae and P. ovale spp., respectively. Prevalence of P. malariae, P. ovale spp., and P. falciparum infections during household visits were 3% (95% CI: 3%-4%), 1% (95% CI: 1%-2%), and 35% (95% CI: 33%-36%), respectively. Non-falciparum malaria was more prevalent in rural and peri-urban vs. urban sites, in school-age children, and among those with P. falciparum co-infection. A crude association was detected between P. malariae and any anemia in the symptomatic clinic population, although this association did not hold when stratified by anemia severity. No crude associations were detected between non-falciparum infection and fever prevalence.

**Conclusions:** *P. falciparum* remains the primary driver of malaria morbidity and mortality in the DRC. However, non-falciparum species also pose an infection risk across sites of varying urbanicity and malaria endemicity within Kinshasa, DRC, particularly among children under 15 years of age. As *P. falciparum* interventions gain traction in high-burden settings like the DRC, continued surveillance and improved understanding of non-falciparum infections are warranted.

## Introduction

Reports of non-falciparum malaria caused by *Plasmodium malariae* and *Plasmodium ovale* spp. have increased across sub-Saharan Africa, where more than 95% of global malaria cases and deaths occur.^1^ *Plasmodium falciparum* is the primary cause of malaria morbidity and mortality in the region. However, molecular surveys have confirmed co-circulating non-falciparum species, and suggest a rise in prevalence in regions where *P. falciparum* has declined.^2–6^ Despite this, our understanding of infection risk and clinical burden posed by non-falciparum species is limited, particularly within regions of high *P. falciparum* transmission.

The Democratic Republic of Congo (DRC) is one of 11 malaria “High Burden, High Impact” countries designated by the World Health Organization (WHO),^7^ indicating a critical need to understand the full landscape of malaria transmission and infection in the country. The DRC harbors the second highest burden of malaria worldwide, accounting for 12% of global malaria cases and 13% of malaria-related deaths as of 2020^1^. Transmission is largely stable and perennial throughout the country, with an estimated 97% of the population living in high malaria transmission regions for eight or more months per year^8–10^. While the majority of the country is characterized as hyper- and holo-endemic for malaria^8,11,12^, transmission can vary within and across provinces due to environmental and geographical factors such as urbanicity, population density, land use, temperature, and rainfall^10,13^. In Kinshasa Province, home to the capital city of Kinshasa, malaria transmission is lower within the central urban areas, and increases among the outer peri-urban and rural zones^10^.

While *P. falciparum* accounts for the vast majority of infections in the DRC, *P. malariae* and *P. ovale* spp. have been documented at low-level prevalences in the DRC^8,14–16^; however, their epidemiology and clinical impact remains poorly understood. Existing non-falciparum prevalence estimates were largely derived from cross-sectional studies, limiting risk assessment, or focused on a singular age group or symptomatic status, which restricts broader generalizability^14,16–22^. The absence of reliable field diagnostics for these less-common infections complicates surveillance efforts and case management. Gold-standard microscopy methods can detect non-falciparum parasites, but insufficient training and specialization ultimately limits the sensitivity and specificity for non-falciparum infections.^23^ Widely-used malaria rapid diagnostic tests (RDTs) in Africa cannot distinguish *P. malariae* or *P. ovale* spp. infections. While most RDTs detect *P. falciparum-*specific histidine-rich protein 2 (HRP2), and some also detect *Plasmodium* lactate dehydrogenase (LDH), no *P. malariae* and *P. ovale* spp.-specific RDTs are currently available. Further, clinical diagnosis of a non-falciparum infection is not feasible in the absence of symptoms attributed specifically to *P. malariae* or *P. ovale* spp. These infections are complicated to detect in malaria-endemic regions such as the DRC where *P. falciparum* co-infection is common, making it difficult to distinguish symptoms caused by these non-falciparum species. *P. malariae* infections have been associated with acute febrile illness and anemia^24–26^ as well as chronic nephrotic syndrome^27,28^. Clinical relevance of *P. falciparum* co-infection with non-falciparum species is also unclear, with posited suppressive effects observed in one study^4^, while another detected severe malaria among mixed-infections^25^. These diagnostic challenges are exacerbated by low parasite densities characteristic of non-falciparum infections, which require the use of sensitive laboratory assays that are not suitable for routine field use.

*Plasmodium malariae* and *P. ovale* spp. infections have occasionally been dismissed as rare infections that are not clinically impactful in high-burden countries like the DRC. However, improved characterization of their epidemiology and clinical implications is needed to determine their proper place in malaria programmatic and surveillance efforts. Leveraging a 34-month longitudinal cohort study conducted across diverse sites in Kinshasa Province, DRC, we sought to determine the epidemiology of *P. malariae* and *P. ovale* spp. infections in the context of high *P. falciparum* transmission. We perform high-throughput, real-time PCR on samples collected from both asymptomatic and symptomatic participants of all ages as part of a large longitudinal study of non-falciparum malaria infection. We estimate incidence, risk factors, and clinical features associated with *P. malariae* and *P. ovale* spp. infections over time, and as compared to *P. falciparum* infections.

## Methods

### Study design

This study leverages data and samples collected during a longitudinal cohort study of malaria transmission across seven sites with varying urbanicity and malaria endemicity in Kinshasa Province, DRC between 2015-2017. Detailed methods for site and household sampling have been previously described.^29,30^ In brief, households were selected at random within villages, and household members were enrolled into the study following screening for eligibility criteria and provision of informed consent, or assent for minors. Participants were followed prospectively through biannual household visits (‘survey-based population’; asymptomatic active surveillance), and at visits to local study health facilities, as-needed, for presentation of fever or other malaria symptoms (“clinic-based subpopulation”; symptomatic passive surveillance) (**Fig 1**). Household surveys were conducted between Feb. 2015 and Oct. 2016 (20 months of active surveillance), with baseline and second follow-up surveys largely occurring during the rainy season, and first and third (final) follow-up surveys occurring during the dry season. Symptomatic clinic visits continued beyond the completion of household surveys, through Dec. 2017, for 34 months of passive surveillance).

**Fig 1.**
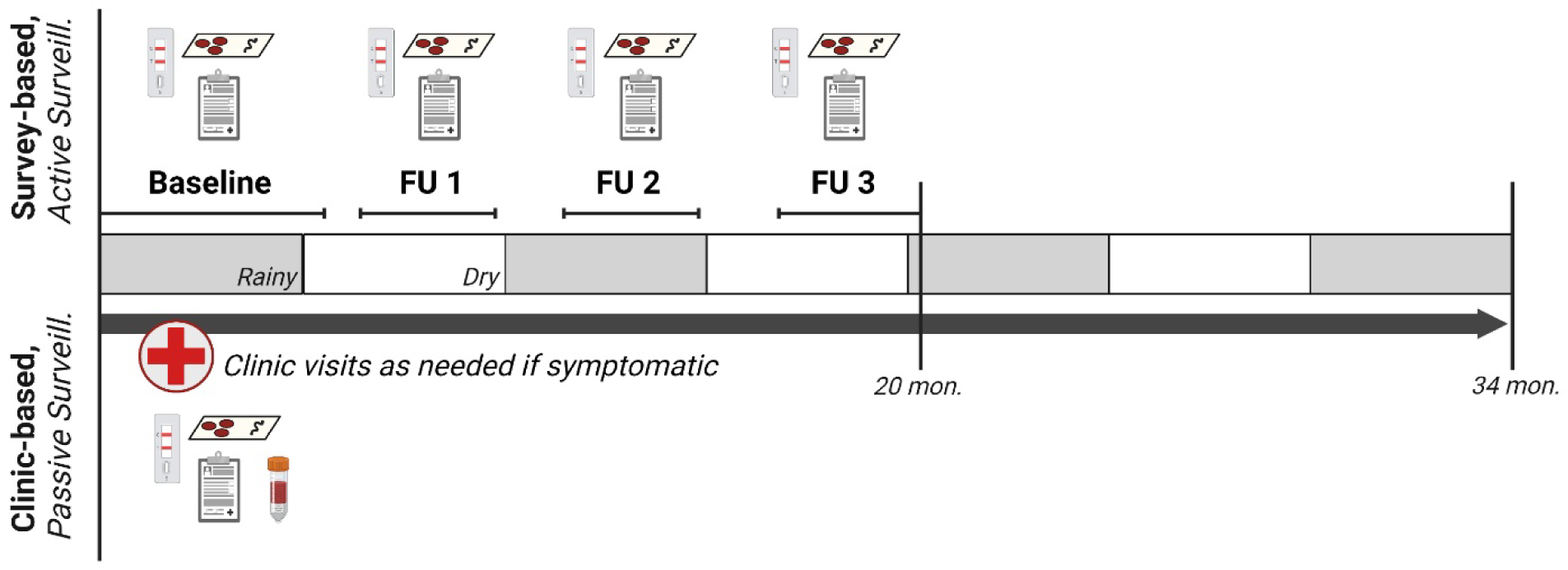
Study design overview^31^. Bi-annual household study visits occurred at baseline and three follow-ups over a 20-month longitudinal period in which household demographic and health surveys were collected, malaria rapid diagnostic tests were performed, and dried blood spots were collected. Participants who developed fever or other malaria symptoms during follow-up visited study health clinics, where clinical questionnaires, malaria rapid diagnostic tests, a dried blood spot, and hemoglobin testing were performed. Study participants completing household surveys were included in the survey population; those who also visited the clinic for malaria symptoms were included in the clinic sub-population.

At baseline and follow-up surveys, household-level and individual-level questionnaires were collected to ascertain demographic information, household characteristics (e.g., housing materials, possessions for wealth indicators, bed net ownership), health status and clinical data (e.g., malaria diagnostic history, recent symptoms, treatment use). Participants were screened for malaria infection at each visit using a combination RDT detecting *P. falciparum*-specific HRP2 and pan-*plasmodium* lactate dehydrogenase (LDH) antigens (SD Bioline Ag P.f./Pan RDT [05FK60], Alere, Gyeonggi-do, Republic of Korea). Those positive by RDT (defined as a positive result for either antigen) were referred to study clinics for antimalarial treatment. Where consented, dried blood spot (DBS) samples were also collected at each visit using Whatman 3MM filter paper (Fisher Scientific, Fair Lawn, NJ USA), and were stored with desiccant at −20°C for future molecular testing.

Brief clinical questionnaires were collected from participants at each symptomatic clinic visit throughout the study, alongside malaria RDTs, DBS samples, and hemoglobin testing. Participants were treated at health facilities according to local clinical judgment, as needed, including artemisinin-based combination therapy (ACT) for malaria infection, or referral for blood transfusion if severely anemic.

### Plasmodium species determination

Samples underwent molecular testing at the University of North Carolina Chapel Hill to distinguish *Plasmodium* species. DNA was extracted from DBS samples using Chelex and saponin^32^. *P. falciparum* parasitemia was identified using a quantitative real-time duplex polymerase chain reaction (PCR) assay targeting the lactate dehydrogenase (*pfldh*) and human beta-tubulin genes, as previously described.^29,32,33^ We tested samples for *P. malariae* and *P. ovale* spp. parasitemia using a semi-quantitative, duplex real-time PCR assay targeting the 18S ribosomal subunit (**S1 Table**).^34^ A single replicate of non-falciparum PCR assays were run on samples and considered positive if amplification occurred at cycle thresholds (Cts) <40. *P. falciparum* samples were run in duplicate and were considered positive if at least one sample amplified at a Ct <38, or both amplified between Cts ≥38 and <40, as previously described.^35^ Semi-quantitative *P. malariae* and *P. ovale* spp. parasite densities were estimated from standard curves made using serially-diluted 18S rRNA plasmid DNA (BEI Resources: #MRA-179 and #MRA-180), assuming six plasmid copies per parasite genome.^36^ Non-falciparum parasitemias were then multiplied by 4.0 to account for a 4-fold dilution of blood from three DBS punches^37^, improving comparability with *P. falciparum* parasitemia quantification. DNA samples with evidence of dehydration were rehydrated prior to PCR testing using 50µL of molecular-grade water; rehydrated samples were excluded from parasite density analyses. Negative (water) controls were included on all PCR runs.

### Population

All participants with a baseline DBS available for testing were included in the study. The Kinshasa longitudinal cohort study population provided two distinct types of data utilized in this study: 1) The “survey-based” analysis consists of participant survey data and clinical samples collected at active surveillance household visits for all those enrolled, and 2) the “clinic-based” analysis consists of participant survey data and clinical samples collected from the subset of the surveyed population who visited study clinics as-needed when symptomatic (passive surveillance).

Samples derived from all study touchpoints (household surveys + clinic visits) were combined to estimate overall infection burden, and also stratified by population type (survey vs. clinic) to account for differences between their predominantly asymptomatic vs. symptomatic nature. Characteristics between the broader survey population and the nested clinic subpopulation were assessed (**S2 Table**), and weighting was deemed unnecessary as factors driving selection for clinic attendance were expected to be directly related to the probability of the outcome.

### Data analysis

The primary outcomes in this study were *P. malariae, P. ovale* spp., and *P. falciparum* infection confirmed by real-time PCR. Demographic and clinical characteristics were summarized by *Plasmodium* species across study populations, and the incidence and prevalence of infections were estimated across the 34-month study follow-up period. Parasite densities were descriptively summarized by species and infection type (mixed vs. single-species).

Cumulative incidences of *P. malariae, P. ovale* spp., and *P. falciparum* infections were calculated using Kaplan-Meier methods among participants who were PCR-negative at baseline for the *Plasmodium* species of interest. Incidences were estimated as the inverse of the survival function, measuring time (months) to the first detected infection since baseline. Participants in the survey population were censored at the time of their last study household visit. Participants in the clinic sub-population were censored at the end of passive surveillance follow-up, as participants were eligible to continue visiting study health clinics even after the final active follow-up, if symptomatic. Participants dropped out of the at-risk population at the time of their first infection event. We assumed no competing risks for malaria infection, a plausible assumption since malaria infection by one species would not preclude infection by another, and mortality events able to be measured in the study were low (n=26, data not shown). Demographic characteristics associated with incidence of infection were evaluated by log-rank testing of stratified cumulative incidence curves.

Prevalences of *P. malariae, P. ovale* spp., and *P. falciparum* were estimated overall, and by infection type, calculated as the number of infections detected out of the total number of visits in the study. Factors associated with non-falciparum infection prevalences were evaluated using binomial generalized estimating equations (GEEs) to account for repeated testing of participants over time. An exchangeable working correlation matrix was assumed for GEE models, and robust standard errors were estimated for calculation of 95% confidence intervals (CIs). We also assessed the frequency of multiple same-species non-falciparum infections detected within a participant throughout follow-up; however, these were not distinguished between acute re-infection events, or chronic carriage of a prior infection.

Household wealth was computed through adaptation of the Demographic and Health Survey (DHS) method and categorized into quintiles as previously described^30,38^; quintiles were collapsed into three categories for analysis, and wealth at baseline was carried forward for all subsequent study visits. Seasonality was defined by month, classifying October through April as the rainy season. Anemia severity at symptomatic clinic visits was classified according to hemoglobin (Hb) level, following World Health Organization (WHO) categories for age-, sex-, and pregnancy-specific cut-points;^40^ severe anemia was defined as Hb<7.0 g/dL for children <5 years, and Hb<8.0 g/dL for all others. Cases were categorized as any anemia, and comparing moderate to severe with mild or no anemia cases. Fever was defined as an axillary temperature >37.5°C measured at symptomatic clinic visits, or as self-reported fever within the prior week at baseline and follow-up surveys. Missing data were summarized and excluded from statistical analyses.

Dataset construction and cleaning were performed using SAS (version 9.4), and analyses were conducted in R (version 4.0.2) using *tidycmprsk, ggsurvfit,* and *gee* packages. Informed consent, and assent where required, was obtained from all participants or their legal guardians prior to study enrollment and sampling. The study was approved by the Institutional Review Boards at the University of North Carolina at Chapel Hill (IRB#: 14-0489) the University of Iowa (#201701201), and the Kinshasa School of Public Health (ESP/CE/015/014).

## RESULTS

### Study population

A total of 1,591 individuals were enrolled in the parent cohort across 242 households and seven sites in Kinshasa Province, DRC. Among these, 1,565 (98.4%) participants had a baseline DBS sample available for analysis and were included in this study, contributing 5,682 total visits. Participant follow-up by analysis population is displayed in **Fig 2**. In the main survey population, 76% of participants completed all three study follow-ups; loss-to-follow-up differed by regional health area, with a higher proportion observed in peri-urban (28%, n=143) and urban (30%, n=116) health areas compared to rural (18%, n=116). Sixty-seven percent (n=1,050) of those in the main survey study also had ≥1 symptomatic visit during the 34-month passive follow-up period and were included in the clinic-based analysis, comprising 218 (90.1%) of the 242 enrolled households. Participants in the clinic subpopulation had 3,407 total clinic visits, with a median (interquartile range [IQR]; min-max) of 2 (1-4; 1-19) visits per person and 11 (5-22; 1-74) per household.

**Fig 2.**
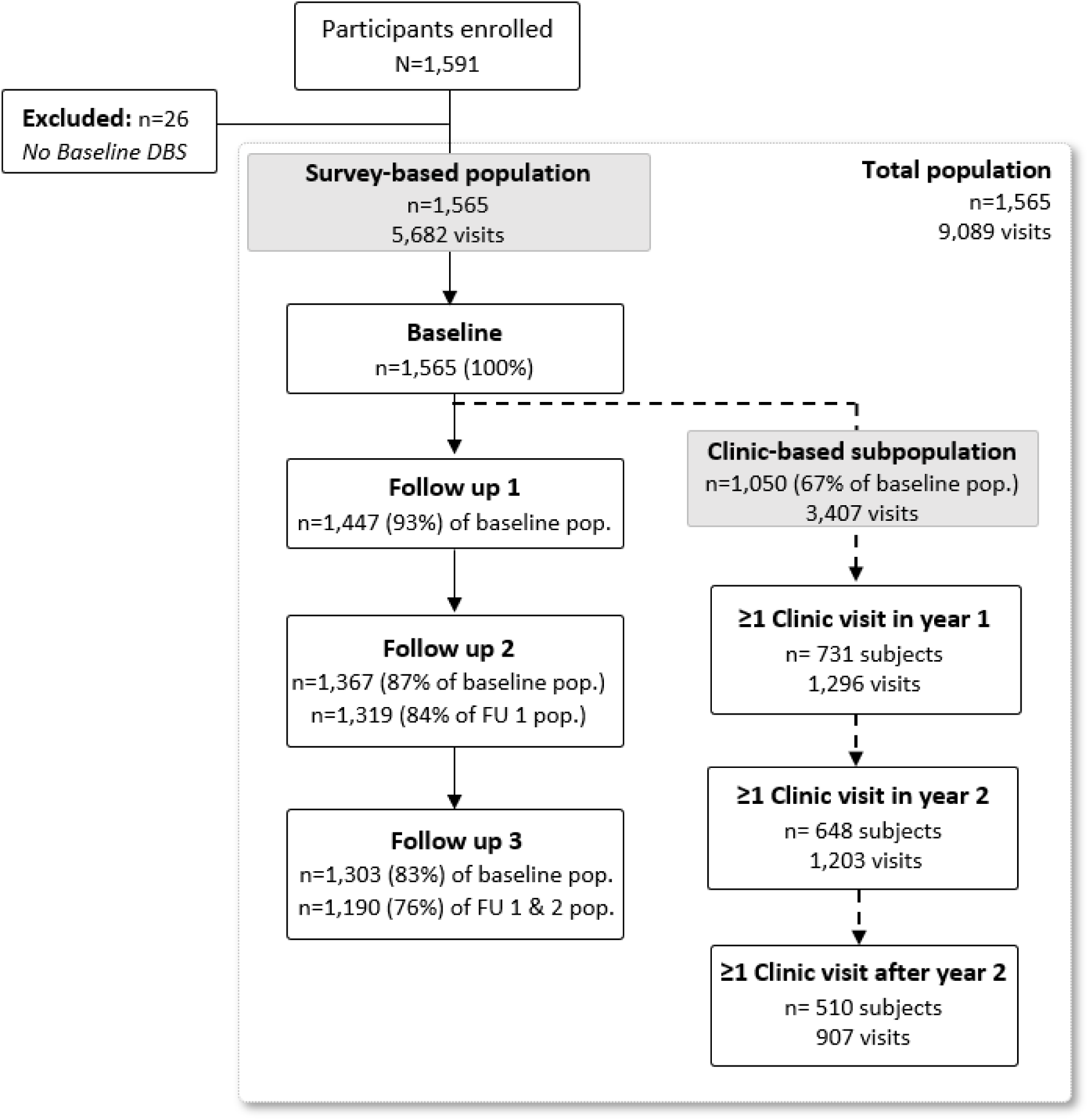
Study population.

Baseline demographic and clinical characteristics for the survey population are summarized in **Table 1**. The median (IQR) age of participants at baseline was 14 (6-31) years, with 19% of participants aged <5 years, and 32% school-aged (between 5-14 years). The majority of participants in the survey population were female (55%), and 42% were living in rural sites. Approximately 73% of the population lived in a household with at least one bed net, although only 45% reported sleeping under a bed net. Twenty-four percent of participants self-reported having a fever in the week prior to baseline, and 25% reported experiencing malaria symptoms in the prior 6 months and taking antimalarials. At the baseline survey, 27% of participants had a positive malaria RDT and 31% were subsequently identified by PCR as having a *P. falciparum* infection, 3% a *P. malariae* infection, and 0.4% a *P. ovale* spp. infection.

**Table 1.** Baseline participant characteristics by species – Survey Population.

| Baseline Participant Characteristics<br>no. (%) | Total Survey Pop.<br>N=1,565 | Baseline Malaria Infection by Species |  |  |
| --- | --- | --- | --- | --- |
|  |  | *P. malariae*<br>PCR Pos.<br>n=47 | *P. ovale* spp.<br>PCR Pos.<br>n=6 | *P. falciparum*<br>PCR Pos.<br>n=484 |
| **Age (years)** |  |  |  |  |
| Median (IQR) | 14 (6–31) | 9 (6–13) | 13 (5–22) | 12 (7–18) |
| <5 | 302 (19.3) | 6 (12.8) | 1 (16.7) | 64 (13.2) |
| 5-14 | 500 (31.9) | 30 (63.8) | 2 (33.3) | 240 (49.6) |
| 15+ | 763 (48.8) | 11 (23.4) | 3 (50.0) | 180 (37.2) |
| **Sex** |  |  |  |  |
| Female | 863 (55.1) | 23 (48.9) | 5 (83.3) | 245 (50.6) |
| Male | 702 (44.9) | 24 (51.1) | 1 (16.7) | 239 (49.4) |
| **Rurality of Household<sup>1</sup>** |  |  |  |  |
| Rural | 663 (42.4) | 28 (59.6) | 6 (100) | 279 (57.6) |
| Peri-urban | 517 (33.0) | 17 (36.2) | 0 (0.0) | 190 (39.3) |
| Urban | 385 (24.6) | 2 (4.3) | 0 (0.0) | 15 (3.1) |
| **Fever in prior week** | 382 (24.4) | 11 (24.4) | 4 (66.7) | 151 (31.2) |
| **RDT+** | 429 (27.4) | 27 (60.0) | 3 (50.0) | 363 (75.0) |
| **Household owns a Bed Net** | 1135 (72.5) | 32 (68.1) | 4 (66.7) | 347 (71.7) |
| **Bed Net Use** | 705 (45.0) | 14 (29.8) | 2 (33.3) | 209 (43.2) |
| **Malaria Sx. ≤ 6 mon.** | 388 (24.8) | 7 (15.6) | 3 (50.0) | 117 (24.2) |
| **Antimalarial use ≤ 6 mon.** | 394 (25.2) | 8 (18.2) | 1 (16.7) | 102 (21.1) |
| **Wealth Category** |  |  |  |  |
| Poorer | 632 (40.4) | 26 (55.3) | 2 (33.3) | 264 (54.5) |
| Average | 311 (19.9) | 10 (21.3) | 2 (33.3) | 107 (22.1) |
| Wealthier | 622 (39.7) | 11 (23.4) | 2 (33.3) | 113 (23.3) |

<sup>1</sup>. Bu health area is classified as rural, Kimpoko health area as peri-urban, and Voix du Peuple as urban.

Baseline characteristics were similar between those also included in the symptomatic clinic subpopulation, and the full survey population (**S2 Table**). The median (IQR) age of participants in the clinic subpopulation was 12 (5-30) years, with 22% aged <5 years, and 34% school-aged (5-14 years). There was also a higher proportion of women (56%) than men in the clinic subpopulation, and higher proportions of participants living in rural (22%) or peri-urban (32%) sites compared to urban sites (46%), as in the broader survey population (**S3 Table**). Participant characteristics across follow-up visits are summarized in **S4** and **S5 Tables**. Overall, 97.0% (n=1,518) and 99.6% (n=1,559) of participants were PCR-negative for a *P. malariae* and *P. ovale* spp. infection at baseline, respectively; 69.1% (n=1,081) were PCR-negative for a *P. falciparum* infection. These PCR-negative participants comprised at-risk populations for malaria incidence estimation.

### Incidence of non-falciparum malaria infections

The estimated 1-year cumulative incidences of *P. malariae* and *P. ovale* spp. infections were 10.6% (95% CI: 8.7%-12.4%) and 6.7% (95% CI: 5.2%-8.2%), respectively, encompassing incident infections detected at survey or clinic visits. Comparatively, 67.2% (95% CI: 63.9%-70.2%) of all participants acquired a *P. falciparum* infection by 1-year (**Fig 3**). Cumulative incidence of *P. malariae* and *P. ovale* spp. was similar within the first year of follow-up, but appears to increase at, and following, the second study follow-up visit around 1-year from baseline, after which the risk of a *P. malariae* infection surpassed that of *P. ovale* spp. Time to the first detected infection was faster for *P. falciparum* than *P. malariae* or *P. ovale* infections throughout the whole study period, with at least half of the total at-risk population experiencing a *P. falciparum* malaria infection within 10 months from baseline. Step-wise increases in incidence depicted in **Fig 3** represent infections detected at periodic household surveys; symptomatic infections detected through passive, clinic-based surveillance filled in gaps between surveys, as indicated by steady inclines between step-ups. The 1-year cumulative incidences of *P. malariae* and *P. ovale* spp. infection detected only at active surveillance household surveys were approximately 4.9% (95% CI: 3.8%-6.1%) and 3.5% (95% CI: 2.5%-4.4%), respectively, including single- and mixed-species infections, whereas the cumulative incidence of any *P. falciparum* infection by 1-year was 43.1% (95% CI: 39.9%-46.1%) within the survey population (**Table 2**).

**Fig 3.**
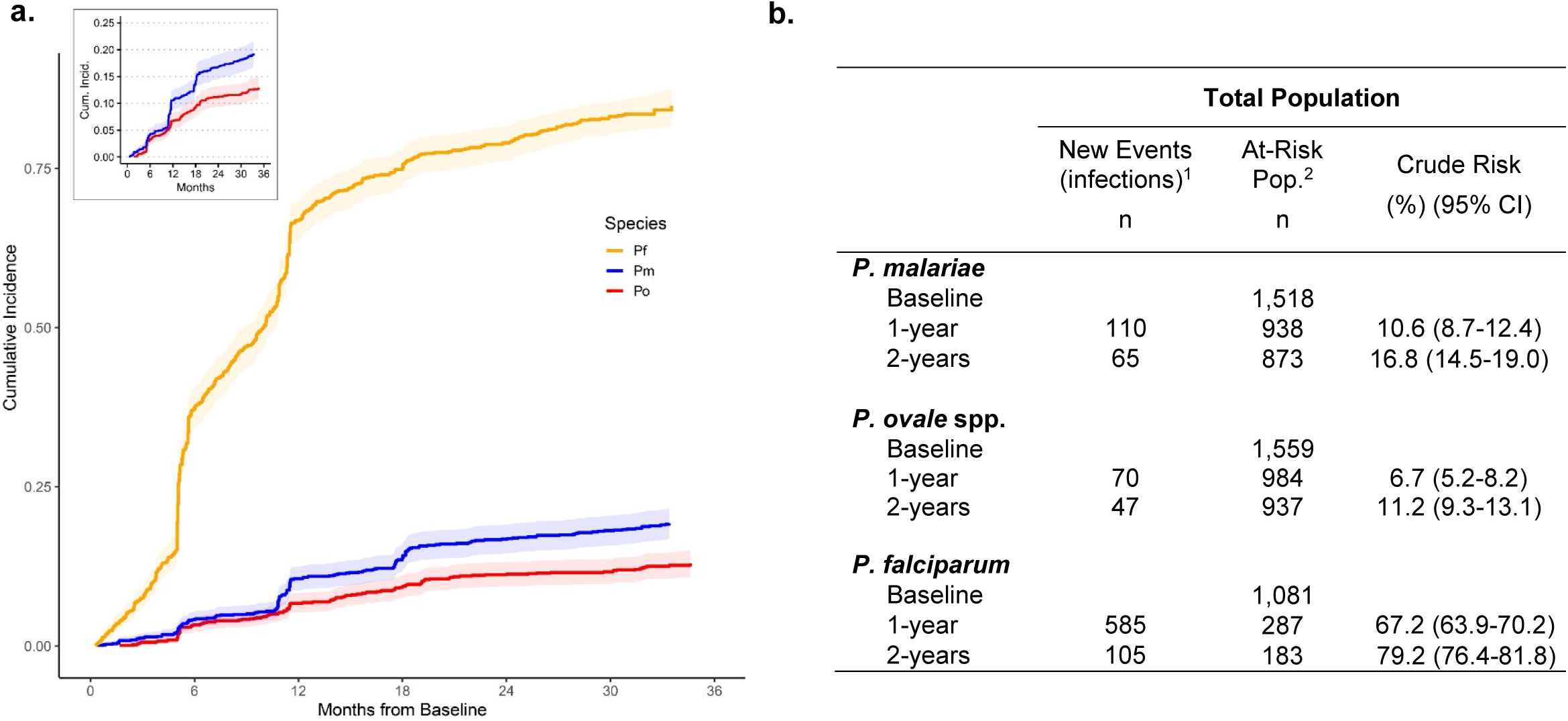
Cumulative incidence of *Plasmodium* infection across 34-months of follow-up in the total study population. **A)** Cumulative incidence curves depict time to the first detected infection among participants negative for each *Plasmodium* species infection at baseline. Shading depicts 95% confidence intervals around incidence estimates. Incidence curves increase in a step-wise fashion at follow-up survey timepoints due to participant-wide screening and case detection, whereas gradual slopes in between shaded follow-up timepoints indicate additional incident events detected through symptomatic presentation to clinics. **B)** 1-year, and 2-year crude risks of incident infection among the total population, by malaria species. New events represent those detected within each time interval; time to the first incident infection only.

**Table 2.** Incidence of non-falciparum infection during household survey visits (asymptomatic) and at clinic visits (symptomatic), among subjects negative for infection at baseline.

|  | Survey Population |  |  | Clinic Sub-population<sup>4</sup> |  |  |
| --- | --- | --- | --- | --- | --- | --- |
|  | New Events (infections)<sup>1</sup><br>n | At-Risk Pop.<sup>2</sup><br>n | Crude Risk (%) (95% CI) | New Events (infections)<sup>1</sup><br>n | At-Risk Pop.<sup>2</sup><br>n | Crude Risk (%) (95% CI) |
| ***P. malariae*** |  |  |  |  |  |  |
| Baseline |  | 1,518 |  |  | 1,002 |  |
| 1-year<sup>3</sup> | 71 | 1,256 | 5.2 (4.0-6.4) | 42 | 910 | 5.0 (3.6-6.4) |
| 2-years | -- | -- | -- | 35 | 875 | 8.7 (6.9-10.4) |
| ***P. ovale* spp.** |  |  |  |  |  |  |
| Baseline |  | 1,559 |  |  | 1,028 |  |
| 1-year<sup>3</sup> | 51 | 1,291 | 3.6 (2.6-4.6) | 30 | 972 | 3.0 (1.9-4.0) |
| 2-years | -- | -- | -- | 34 | 938 | 6.4 (4.9-7.9) |
| ***P. falciparum*** |  |  |  |  |  |  |
| Baseline |  | 1,081 |  |  | 727 |  |
| 1-year<sup>3</sup> | 431 | 685 | 43.4 (40.2-46.4) | 341 | 323 | 51.5 (47.6-55.2) |
| 2-years | -- | -- | -- | 111 | 211 | 68.3 (64.5-71.6) |

<sup>1</sup> Number of incident infections detected in each time interval. Time to the first incident infection only.
<sup>2</sup> At-risk population comprises subjects PCR-negative for each species-specific infection at Baseline.
<sup>3</sup> For the survey-based population, 1-year incidence estimates are approximated as the incidence of infection by the end of Follow-up 2, inclusive of all events through Day 430 from Baseline (mean [SD] days from Baseline to Follow-up 2: *Po/Pm* - 337 [20] days; *Pf* - 331 [21] days).
<sup>4</sup> Includes infections detected only at symptomatic clinic visits.

### Characteristics associated with incidence of non-falciparum and P. falciparum infections

Cumulative incidences of non-falciparum and *P. falciparum* infection stratified by age and sex are depicted in **Fig. 4**. Within the total study population, cumulative incidences of *P. malariae* and *P. ovale* spp. infection were higher among children <5 and school-aged children 5-14 years old compared to participants aged ≥15 years (p<0.001). Time to the first detected *P. ovale* spp. infection was similar between children <5 and 5-14 years throughout follow-up, whereas school-aged children 5-14 years old experienced a faster time to *P. malariae* infection, particularly towards the end of follow-up driven by symptomatic clinic-based infections. The cumulative incidence of *P. falciparum* infection was also higher among children <5 and 5-14 years old, compared to adults aged ≥15, although *P. falciparum* incidences were high (>60%) across all age categories by 1-year (p=0.003). No differences in the cumulative incidence of *P. ovale* spp. and *P. falciparum* infection by sex were detected (p=0.57 and p=0.40, respectively), although *P. malariae* infection incidence was higher among males than females (p=0.02). Incidence of *P. malariae, P. ovale* spp., and *P. falciparum* infections over time was similar between rural and peri-urban settings, but was significantly lower among urban households (p<0.001 across species), with risk of *P. malariae* and *P. ovale* spp. close to zero in urban areas across the full study period encompassing survey and clinic-based infections.

**Figure 4.**
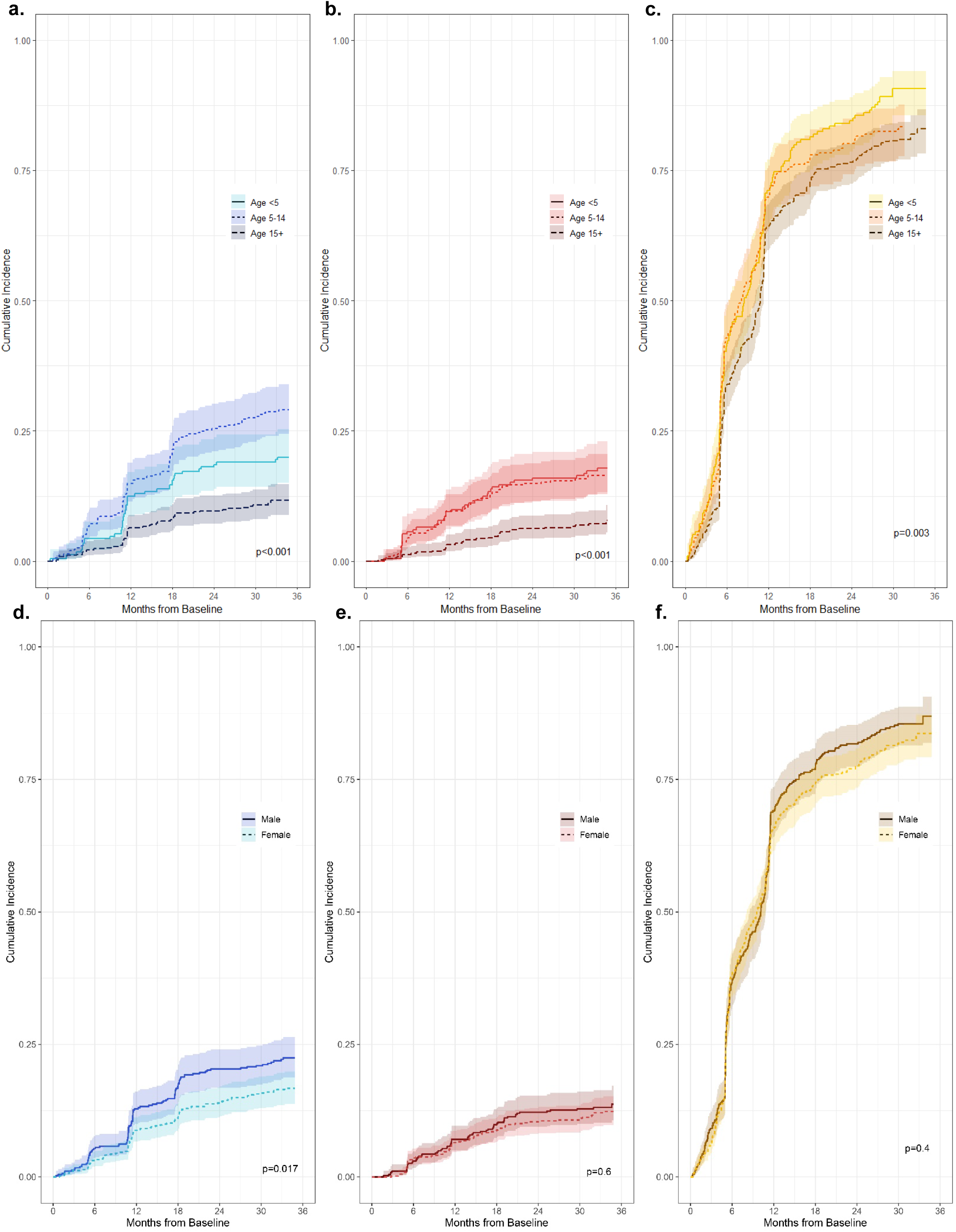
Cumulative incidence curves stratified by participant characteristics: **a-c)** Cumulative incidence of first infection, stratified by age at baseline, for *P. malariae* (a), *P. ovale* spp. (b), and *P. falciparum* (c) species; **d-f)** Cumulative incidence of first infection, stratified by sex, for *P. malariae* (d), *P. ovale* spp. (e), and *P. falciparum* (f).

### Prevalence of non-falciparum and P. falciparum infections

Crude prevalence of non-falciparum infection was lower in household survey visits than symptomatic clinic visits, as expected. Across all household surveys, we observed a crude period prevalence of 3.3% (95% CI: 2.8%-3.8%) for *P. malariae* infection, and 1.4% (95% CI: 1.1%-1.7%) for *P. ovale* spp. infection. Prevalence of *P. falciparum* infection was higher at 34.9% (95% CI: 33.2%-36.5%). The majority of *P. malariae* and *P. ovale* spp. infections in the survey-based population were mixed-species (*P. malariae:* 73.7% [n=137]; *P. ovale* spp.: 78.2% [n=61]), and were predominately co-infected with *P. falciparum*. In contrast, the vast majority of *P. falciparum* infections were single-species (90.5%; n=1,788).

Similar prevalences to those detected in the survey population were observed among the symptomatic clinic subpopulation, for *P. malariae* (4.0% [95% CI: 3.3%-4.7%]) and *P. ovale* spp. (2.8% [95% CI: 2.2%-3.4%]. However, prevalence of *P. falciparum* infection was higher in the symptomatic clinic subpopulation (58.7% [95% CI: 56.5%-60.8%]) compared to the survey population.

*P. malariae* prevalence across household survey visits remained steady throughout follow-up, fluctuating between 3.0% at baseline, 2.4% at follow-up 1, 4.1% at follow-up 2, and 3.7% at the final household survey. Similarly, *P. ovale* spp. prevalence remained relatively low throughout follow-up, with 0.4% of the population infected with *P. ovale* spp. at baseline, increasing to 1.9% and 2.0% at follow-ups 1 and 2, and ending at a 1.4% prevalence by the final follow-up. The timeline of non-falciparum infections by participant throughout the study are depicted in **S2 Fig.** Multiple non-falciparum infections of the same species were detected over time in 23.5% (n=58/247) of all participants who had a *P. malariae* infection, and in 21.6% (n=30/139) of those with a *P. ovale* spp. infection during the 34-month study period. Three participants each had a *P. malariae* infection detected 4 times throughout follow-up. In comparison, *P. falciparum* infections re-occurred frequently, with 75.2% (n=914/1216) of those who had a *P. falciparum* infection having more than one during follow-up, and 22.4% (n=272/1216) having ≥5 *P. falciparum* infections detected.

Children had non-falciparum infections more frequently than adults. Prevalence of *P. malariae* infection in the survey-based population was highest among school-aged children aged 5-14 years at 5.6% (95% CI: 4.4-6.7%), though still low overall, followed by children <5 years (3.4% [95% CI: 2.1-4.7%]), and lowest among those aged ≥15 (1.7% [95% CI: 1.2%-2.2%]). Conversely, children <5 years experienced a slightly higher prevalence of *P. ovale* spp. infection compared to school-aged children (2.3% [95% CI: 1.3%-3.3%] vs. 1.9% [95% CI: 1.2%-2.7%]), although prevalences across both age groups were low overall, and lower than *P. malariae* prevalence in these age groups; 0.7% (95% CI: 0.4% - 1.0%) of adults ≥15 years had a *P. ovale* spp. infection at study survey visits.

Factors associated with the prevalence of non-falciparum infections are shown in **Fig. 5** and contrasted with *P. falciparum* prevalence differences in **S1 Fig**. Within the survey-based population, *P. falciparum* coinfection was associated with an increased prevalence of both *P. malariae* and *P. ovale* spp. infections. In comparison to those with average wealth, higher wealth was associated with a decreased prevalence of *P. malariae* and *P. ovale* spp. infection. School-aged children 5-14 years old were associated with a higher prevalence of *P. malariae* and *P. ovale* spp. infection as compared to participants 15 and older, and also as compared to children aged <5 for *P. malariae*. Similarly, in the clinic-based analysis, school-aged children were also associated with an increased prevalence of *P. malariae* and *P. ovale* spp., infections, compared to adults ≥15 and older; however, prevalence was similar between children <5 years and adults ≥15 and older in this symptomatic population. Likewise, no associations were observed with *P. falciparum* co-infection for either non-falciparum species.

**Fig 5.**
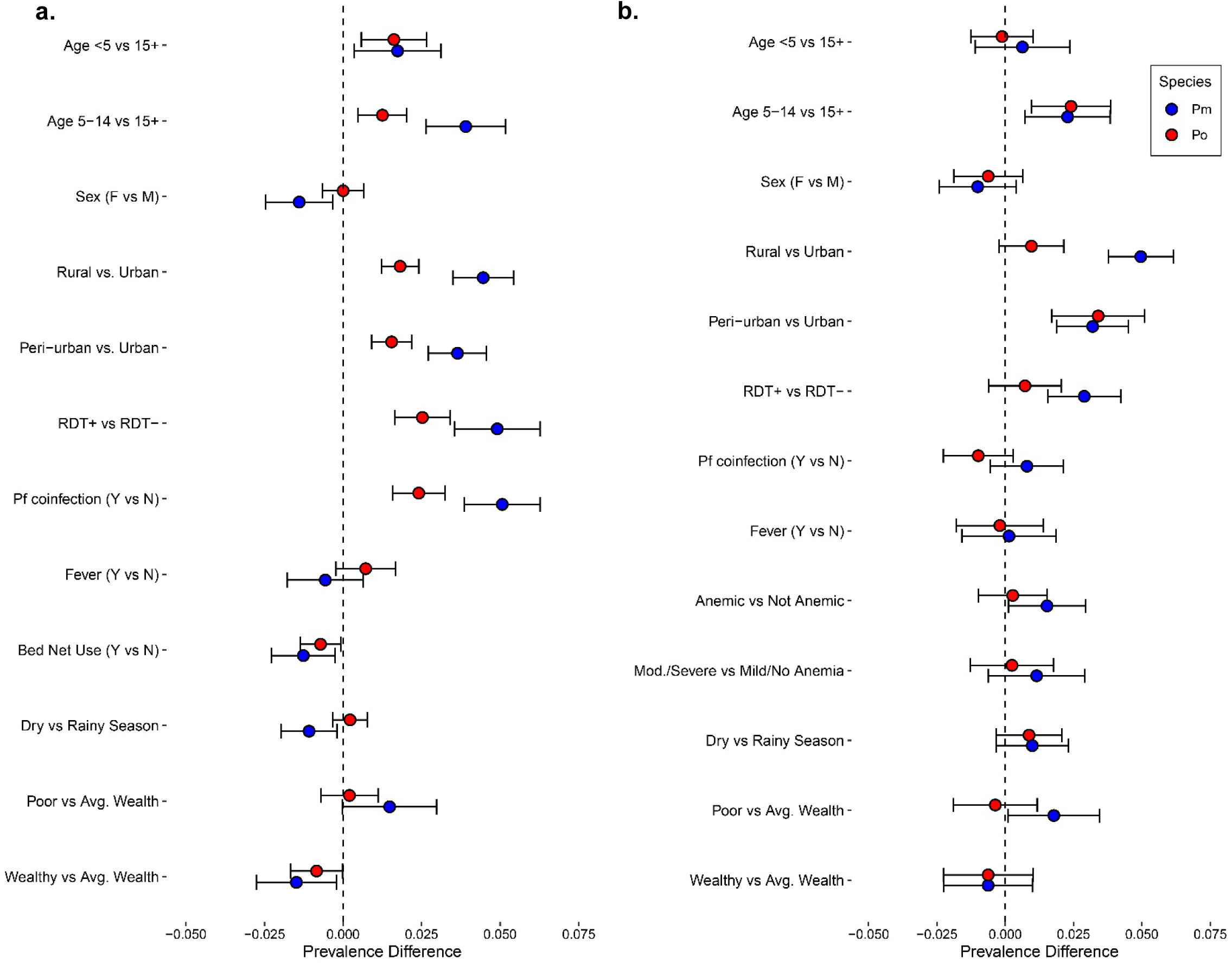
Factors associated with *P. malariae* and *P. ovale* spp. infection prevalence, stratified by study population. **a)** Factors associated with non-falciparum infection prevalence at survey visits (baseline and three follow-up surveys). **b)** Factors associated with non-falciparum prevalence at symptomatic clinic visits across follow-up (i.e., symptomatic cases).

Prevalence of self-reported fever in the prior week was similar for *P. malariae* (PD: −0.006 [95% CI:-0.018-0.006]) and *P. ovale spp.* infections (PD: 0.007 [95% CI: −0.002-0.017]), as compared to no malaria infection among the survey population (**Fig 5**). In the symptomatic clinic subpopulation, concurrent anemia of any severity was associated with *P. malariae* infection prevalence (PD: 0.015 [95% CI: 0.001-0.029]), although associations were no longer significant when stratifying by anemia severity, comparing moderate-to-severe cases vs. mild or no anemia for *P. malariae* (PD: 0.012 [95% CI: −0.006-0.029]), or for *P. ovale* spp. (PD: 0.002 [95% CI: −0.013-0.018].

### Differences in parasite densities by species and during co-infection

Estimated median (IQR) parasite densities were low (<50 p/µL) among all non-falciparum infections in the study (*P. malariae*: 25.7 [7.7-119] p/µL; *P. ovale*: 10.2 [2.7-47.4] p/µL), while *P. falciparum* parasite densities in the total population were higher at 267 (18.8-4,526) p/µL. Non-falciparum parasite densities remained low when stratified by asymptomatic survey population (*P. malariae*: 22.4 [8.5-72.2] parasites[p]/µL; *P. ovale* spp.: 5.8 [2.0-28.0] p/µL), vs. symptomatic clinic sub-population (*P. malariae*: 36.5 [4.7-182] p/µL; *P. ovale*: 17.7 [4.6-65.8] p/µL). Parasite density distributions were slightly higher among *P. malariae* infections than *P. ovale* spp. infections. We did not detect any differences in estimated *P. malariae* or *P. ovale* spp. parasite density distributions between mixed- vs. single-species *P. malariae* or *P. ovale* spp. infections in the total population (*P. malariae:* p=0.071; *P. ovale*: p=0.465), or across specific population types. Parasite densities are summarized in **S5 Table**; samples rehydrated during molecular analysis were excluded from analysis (n=591 total).

Interestingly, differences in the distribution of *P. falciparum* parasite densities were observed between single-species *P. falciparum* infections vs. mixed-species infections with *P. malariae,* but not mixed *P. ovale* spp. (**Fig 6**). A higher median (IQR) parasite density was observed within mixed *P. falciparum-P. malariae* co-infections (127 [25-496] p/µL vs. 47 [7-311] p/µL; p=0.001) and *P. falciparum-P. ovale* co-infections (122 [42-486] p/µL vs. 47 [7-311] p/µL; p=0.001) as compared to single-species *P. falciparum* infections among the asymptomatic survey population. However, a lower median *P. falciparum* parasite density was observed within mixed *P. falciparum-P. malariae* infections compared to *P. falciparum* single-species infections in the clinic-based sub-population (288 [57-4,319] p/µL vs. 2897 [127-18,058 p/µL; p<0.001); this association was not observed for *P. falciparum-P. ovale* co-infections (1,710 [165-13,082] p/µL vs. 2,897 [127-18,058] p/µL; p=0.596), although the sample size for mixed *P. falciparum-P. ovale* co-infections was smaller than for single-species *P. falciparum* infections.

**Fig 6.**
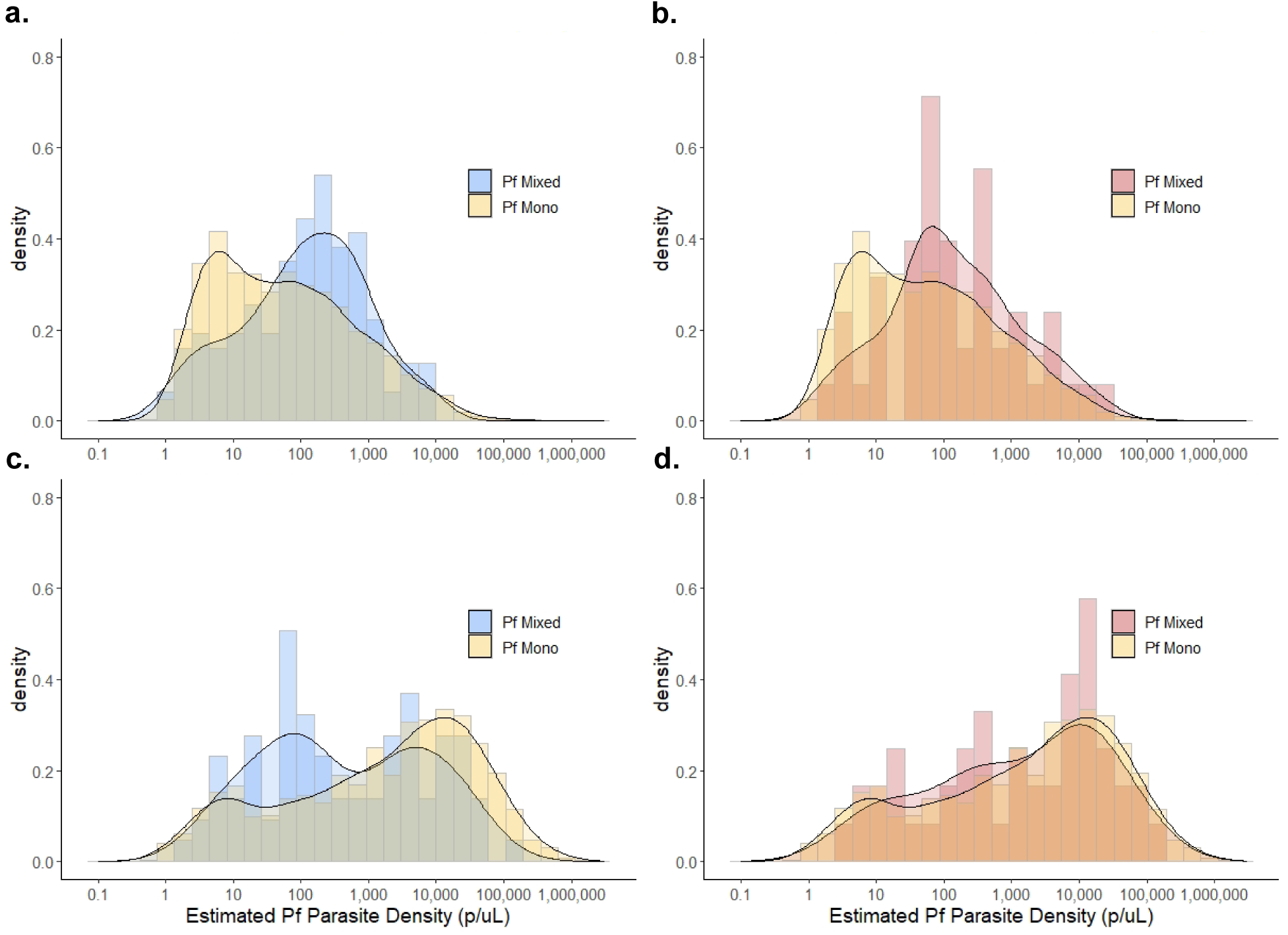
Distributions of *P. falciparum* parasitemia by type of non-falciparum mixed infection vs. *P. falciparum* mono infections, stratified by population. Household survey-based results for *P. falciparum* parasitemia distributions comparing *P. falciparum* mono infections vs. co-infection with a) *P. malariae* and b) *P. ovale* spp. Clinic sub-population *P. falciparum* parasitemia distributions comparing *P. falciparum* mono infections vs. co-infection with c) *P. malariae* and d) *P. ovale* spp. Density distributions are displayed to illustrate trends in overall distributions accounting for variation in sample sizes across groups.

## Discussion

Non-falciparum malaria infection was common in this study, affecting children and adults across a range of *P. falciparum* transmission intensities in Kinshasa Province, DRC. In the largest longitudinal study of non-falciparum malaria conducted to-date in the DRC, we observed an 11% cumulative incidence of *P. malariae* infection, and 7% incidence of *P. ovale* spp. infection within one-year, including both asymptomatic and symptomatic infections. Over two-thirds of the study population in this high malaria burdened country had an incident *P. falciparum* infection within one year, confirming the central importance of *P. falciparum* in the DRC. Over 70% of the non-falciparum infections detected occurred as co-infections with *P. falciparum*. Though *P. falciparum* was the dominant species, we detected a considerable risk of infection by a non-falciparum species, particularly for children under 15 years of age. These findings underscore the need to consider these neglected species in areas of high *P. falciparum* transmission.

Ten percent of asymptomatic household survey participants had at least one *P. malariae* infection during the study, and 7% had at least one *P. ovale* spp. infection, amounting to 3% and 1% *P. malariae* and *P. ovale* spp. infection prevalence at study visits across follow-up, including both mixed- and single-species infections. Similar prevalences were observed among symptomatic individuals. Prevalences generally align with previous estimates of non-falciparum infections in the DRC derived from cross-sectional studies^14,16–22,41^. Infection prevalences were stable across the 34-month follow-up, differing from recent studies in neighboring countries which have detected increases in *P. malariae* and *P. ovale* infection prevalence over time, particularly as *P. falciparum* prevalences have started to decline^3,4,6^. The high proportion of *P. malariae*-infected participants who experienced multiple infections in the study (24%) is notable given that this species is known to recrudesce. Chronic *P. malariae* infection may be associated with deleterious clinical outcomes, such as severe anemia and chronic nephrotic syndrome^25,26,42^. Though we could not distinguish whether multiple *P. malariae* infections were chronic occurrences or acute re-infection events, future work may allow us to differentiate these infection types. Such work is necessary given the dearth of evidence on the commonality and timing of non-falciparum infection recrudescence, as well as reinfection rates for non-falciparum species in regions highly endemic for *P. falciparum*.

Populations at risk for *P. malariae* and *P. ovale* spp. infection were similar but not identical to those at risk for *P. falciparum.* School-aged children were found to have higher prevalences of *P. malariae* infection than children under 5 years of age, following similar age-related infection associations with *P. falciparum* that have been previously established in the region^30,43^. Dissimilar from *P. falciparum* infection patterns, symptomatic children under 5 had comparable *P. malariae* infection prevalences as those experienced by symptomatic adults aged 15 and older. All *Plasmodium* species were generally more common in rural and peri-urban sites, and less common in the wealthiest households.

We observed a crude association between *P. malariae* infection prevalence and anemia of any severity in the symptomatic clinic subpopulation, which translated to a 1.5% absolute increase in prevalence compared to those without anemia. Although this association was no longer significant when stratifying by anemia severity, and was not detected among *P. ovale* spp. infections in this study. Similarly, no association with non-falciparum infections and fever was detected within the survey- or clinic-based analyses, though misclassification of fever is possible in the setting of fever-reducing medicines taken prior to clinic visits, for example. Whether fever-reducing medicines were taken was not captured in the study. Prior studies have posited a protective association between non-falciparum-*P. falciparum* co-infections and lower prevalences of clinical malaria outcomes as compared to single-species *P. falciparum* infections ^44,45^, although this has not been consistently observed across studies^46,47^. We did not assess causal effects of non-falciparum infection on clinical outcomes in our study (though no crude associations were detected), including possible attenuation of symptomatic *P. falciparum* due to mixed-species infections.

Interestingly, *P. falciparum* parasite densities were higher in mixed infections with *P. malariae* or *P. ovale* spp. in the household survey-based, but not clinic-based, analysis. One possible explanation is that *P. falciparum* increases its virulence to compete with non-falciparum species within the asymptomatic host, though our data are insufficient to evaluate this hypothesis. Non-falciparum parasite densities observed in this study were low, as is characteristic of *P. malariae* and *P. ovale* spp. infections. We did not identify any difference in non-falciparum parasite density between mixed and single-species non-falciparum infections.

Our incidence and prevalence estimates should be considered lower bounds of the true incidence and prevalence in the study catchment area owing to several limitations of the study. First, while loss to follow-up was low overall in the study, missed study visits, early loss to follow-up, and differences in study health clinic attendance across sites may have introduced selection bias into the study. Participants may have elected to visit alternative health facilities for malaria care instead of presenting to study clinics. However, as all participants were eligible for free treatment at study clinics, the clinic sub-population is likely to be representative of the overall study population. Second, our duplex PCR assay detected low density infections but would likely miss those with very low densities. Third, the biannual frequency of household surveys missed infections that occurred and resolved between study visits. Similarly, while the longitudinal design provides helpful insight into risk of infection surrounding these species, even 34-months of follow-up is likely insufficient to detect changes in the prevalence of these less-common species over time in the DRC, where gains in malaria control have not been achieved as in other settings. Fourth, some misclassification of seasonality may be present, as local classifications for seasonality by month, rather than rainfall amounts, were used to define rainy seasons. Finally, our observational study design is not immune from the risk of unmeasured confounding, although no causal relationships were assessed in this study as risk factors depict crude associations, and the large sample size improves confidence in our conclusions.

This study provides insight into the epidemiology of non-falciparum species in a region heavily affected by *P. falciparum.* Though less common and impactful than *P. falciparum, P. malariae* and *P. ovale* spp. infections occurred frequently and were often detected within symptomatic cases in this large longitudinal study in the DRC. Malaria research and control efforts focused on *P. falciparum* should consider these neglected species, particularly in school-aged children and rural communities.

## DATA AVAILABILITY

Data will be made publicly available through the Carolina Digital Repository.

## Supporting information

STROBE Checklist

## Data Availability

Data will be made publicly available through the Carolina Digital Repository.

## Acknowledgements

We thank the study team for conducting household surveys and field sampling, as well as all study participants for their time and engagement throughout multiple years of follow-up. We also thank the late Prof. Steven Meshnick for his leadership in establishing the parent study and for mentorship to DRC- and US-based co-authors. Lastly, we thank Seungwon Kim at the University of Iowa for their calculation of wealth scores by principal component analysis (PCA) provided for this analysis. The following reagents were obtained through BEI Resources, NIAID, NIH: Diagnostic Plasmid Containing the Small Subunit Ribosomal RNA Gene (18S) from *Plasmodium malariae,* MRA-179, and *Plasmodium ovale*, MRA-180, contributed by Peter A. Zimmerman. The following reagent was obtained through BEI Resources, NIAID, NIH: *Plasmodium falciparum*, Strain 3D7, MRA-102, contributed by Daniel J. Carucci.

## Author Contributions

- Conception and study design: RS, KB, JBP
- Laboratory work: RS, MS, KLT
- Data collection and cleaning: RS, KB, MMK, NM, MN, GK, JA, KLT, WMS, VG, TN, JBP
- Data analysis and interpretation: RS, KB, VG, ME, MC, JJJ, AT, JTL, JBP
- Drafting manuscript: RS, KB, JBP
- Review and approval of manuscript: all authors

## REFERENCES

1. World Malaria Report 2021. Genève, Switzerland: World Health Organization; 2021.

2. Oboh MA, Oyebola KM, Idowu ET, Badiane AS, Otubanjo OA, Ndiaye D. Rising report of Plasmodium vivax in sub-Saharan Africa: Implications for malaria elimination agenda. Scientific African. 2020;10:e00596.

3. Hawadak J, Dongang Nana RR, Singh V. Global trend of Plasmodium malariae and Plasmodium ovale spp. malaria infections in the last two decades (2000-2020): a systematic review and meta-analysis. Parasit Vectors. 2021;14(1):297.

4. Akala HM, Watson OJ, Mitei KK, et al. Plasmodium interspecies interactions during a period of increasing prevalence of Plasmodium ovale in symptomatic individuals seeking treatment: an observational study. Lancet Microbe. 2021;2(4):e141–e150.

5. Twohig KA, Pfeffer DA, Baird JK, et al. Growing evidence of Plasmodium vivax across malaria-endemic Africa. PLoS Negl Trop Dis. 2019;13(1):e0007140.

6. Yman V, Wandell G, Mutemi DD, et al. Persistent transmission of Plasmodium malariae and Plasmodium ovale species in an area of declining Plasmodium falciparum transmission in eastern Tanzania. PLoS Negl Trop Dis. 2019;13(5):e0007414.

7. World Health Organization. High burden to high impact: a targeted malaria response. 2019. https://apps-who-int.libproxy.lib.unc.edu/iris/bitstream/handle/10665/275868/WHO-CDS-GMP-2018.25-eng.pdf.

8. PNLP, KSPH, Swiss KSPH, INRB and INFORM. An Epidemiological Profile of Malaria in the Democratic Republic of Congo. A Report Prepared for the Federal Ministry of Health, Democratic Republic of Congo, the Roll Back Malaria Partnership and the Department for International Development, UK.; 2014. http://www.inform-malaria.org/wp-content/uploads/2015/03/DRC-Epidemiological-Report-120914.pdf.

9. Fy MOP. Democratic Republic of Congo (DRC).

10. Ferrari G, Ntuku HM, Schmidlin S, Diboulo E, Tshefu AK, Lengeler C. A malaria risk map of Kinshasa, Democratic Republic of Congo. Malar J. 2016;15:27.

11. Nardini L, Hunt RH, Dahan-Moss YL, et al. Malaria vectors in the Democratic Republic of the Congo: the mechanisms that confer insecticide resistance in Anopheles gambiae and Anopheles funestus. Malar J. 2017;16(1):448.

12. U.S. President’s Malaria Initiative. Democratic Republic of the Congo Malaria Operational Plan FY 2023. https://d1u4sg1s9ptc4z.cloudfront.net/uploads/2023/01/FY-2023-DR-Congo-MOP.pdf.

13. Stresman GH. Beyond temperature and precipitation: ecological risk factors that modify malaria transmission. Acta Trop. 2010;116(3):167–172.

14. Doctor SM, Liu Y, Anderson OG, et al. Low prevalence of Plasmodium malariae and Plasmodium ovale mono-infections among children in the Democratic Republic of the Congo: a population-based, cross-sectional study. Malar J. 2016;15:350.

15. Messina JP, Taylor SM, Meshnick SR, et al. Population, behavioural and environmental drivers of malaria prevalence in the Democratic Republic of Congo. Malar J. 2011;10:161.

16. Taylor SM, Messina JP, Hand CC, et al. Molecular malaria epidemiology: mapping and burden estimates for the Democratic Republic of the Congo, 2007. PLoS One. 2011;6(1):e16420.

17. Mitchell CL, Brazeau NF, Keeler C, et al. Under the Radar: Epidemiology of Plasmodium ovale in the Democratic Republic of the Congo. J Infect Dis. 2021;223(6):1005–1014.

18. Kiyonga Aimeé K, Lengu TB, Nsibu CN, Umesumbu SE, Ngoyi DM, Chen T. Molecular detection and species identification of Plasmodium spp. infection in adults in the Democratic Republic of Congo: A population-based study. PLoS One. 2020;15(11):e0242713.

19. Mvumbi DM, Bobanga TL, Melin P, et al. High Prevalence of *Plasmodium falciparum* Infection in Asymptomatic Individuals from the Democratic Republic of the Congo. Malar Res Treat. 2016;2016:5405802.

20. Nundu SS, Culleton R, Simpson SV, et al. Malaria parasite species composition of Plasmodium infections among asymptomatic and symptomatic school-age children in rural and urban areas of Kinshasa, Democratic Republic of Congo. Malar J. 2021;20(1):389.

21. Kavunga-Membo H, Ilombe G, Masumu J, et al. Molecular identification of Plasmodium species in symptomatic children of Democratic Republic of Congo. Malar J. 2018;17(1):334.

22. Parr JB, Kieto E, Phanzu F, et al. Analysis of false-negative rapid diagnostic tests for symptomatic malaria in the Democratic Republic of the Congo. Sci Rep. 2021;11(1):6495.

23. Fançony C, Sebastião YV, Pires JE, Gamboa D, Nery SV. Performance of microscopy and RDTs in the context of a malaria prevalence survey in Angola: a comparison using PCR as the gold standard. Malar J. 2013;12:284.

24. Ayo D, Odongo B, Omara J, et al. Plasmodium malariae infections as a cause of febrile disease in an area of high Plasmodium falciparum transmission intensity in Eastern Uganda. Malar J. 2021;20(1):425.

25. Douglas NM, Lampah DA, Kenangalem E, et al. Major burden of severe anemia from non-falciparum malaria species in Southern Papua: a hospital-based surveillance study. PLoS Med. 2013;10(12):e1001575; discussion e1001575.

26. Langford S, Douglas NM, Lampah DA, et al. Plasmodium malariae Infection Associated with a High Burden of Anemia: A Hospital-Based Surveillance Study. PLoS Negl Trop Dis. 2015;9(12):e0004195.

27. Mueller I, Zimmerman PA, Reeder JC. Plasmodium malariae and Plasmodium ovale--the “bashful” malaria parasites. Trends Parasitol. 2007;23(6):278–283.

28. Lo E, Nguyen K, Nguyen J, et al. Plasmodium malariae Prevalence and csp Gene Diversity, Kenya, 2014 and 2015. Emerg Infect Dis. 2017;23(4):601–610.

29. Mwandagalirwa MK, Levitz L, Thwai KL, et al. Individual and household characteristics of persons with Plasmodium falciparum malaria in sites with varying endemicities in Kinshasa Province, Democratic Republic of the Congo. Malar J. 2017;16(1):456.

30. Carrel M, Kim S, Mwandagalirwa MK, et al. Individual, household and neighborhood risk factors for malaria in the Democratic Republic of the Congo support new approaches to programmatic intervention. Health Place. 2021;70:102581.

31. Figure created with Biorender.com.

32. Plowe CV, Djimde A, Bouare M, Doumbo O, Wellems TE. Pyrimethamine and proguanil resistance-conferring mutations in Plasmodium falciparum dihydrofolate reductase: polymerase chain reaction methods for surveillance in Africa. Am J Trop Med Hyg. 1995;52(6):565–568.

33. Pickard AL, Wongsrichanalai C, Purfield A, et al. Resistance to antimalarials in Southeast Asia and genetic polymorphisms in pfmdr1. Antimicrob Agents Chemother. 2003;47(8):2418–2423.

34. Rougemont M, Van Saanen M, Sahli R, Hinrikson HP, Bille J, Jaton K. Detection of four Plasmodium species in blood from humans by 18S rRNA gene subunit-based and species-specific real-time PCR assays. J Clin Microbiol. 2004;42(12):5636–5643.

35. Doctor SM, Liu Y, Whitesell A, et al. Malaria surveillance in the Democratic Republic of the Congo: comparison of microscopy, PCR, and rapid diagnostic test. Diagn Microbiol Infect Dis. 2016;85(1):16–18.

36. Gumbo A, Topazian HM, Mwanza A, et al. Occurrence and Distribution of Nonfalciparum Malaria Parasite Species Among Adolescents and Adults in Malawi. J Infect Dis. 2022;225(2):257–268.

37. Hewawasam E, Liu G, Jeffery DW, Gibson RA, Muhlhausler BS. Estimation of the volume of blood in a small disc punched from a dried blood spot card. Eur J Lipid Sci Technol. 2018;120(3):1700362.

38. Rutstein SO, Johnson K. The DHS Wealth Index.; 2004. http://dhsprogram.com/publications/pdf/CR6/CR6.pdf. Accessed September 22, 2022.

39. Shuttle Radar Topography Mission. https://www2.jpl.nasa.gov/srtm/. Accessed January 9, 2023.

40. World Health Organization. Haemoglobin concentrations for the diagnosis of anaemia and assessment of severity. https://apps.who.int/iris/bitstream/handle/10665/85839/WHO_NMH_NHD_MNM_11.1_eng.pdf.

41. Brazeau NF, Mitchell CL, Morgan AP, et al. The epidemiology of Plasmodium vivax among adults in the Democratic Republic of the Congo. Nat Commun. 2021;12(1):4169.

42. Gilles HM, Hendrickse RG. Nephrosis in Nigerian children. Role of Plasmodium malariae, and effect of antimalarial treatment. Br Med J. 1963;2(5348):27–31.

43. Deutsch-Feldman M, Parr JB, Keeler C, et al. The Burden of Malaria in the Democratic Republic of the Congo. doi:10.1093/infdis/jiaa650#supplementary-data

44. Bruce MC, Macheso A, Kelly-Hope LA, Nkhoma S, McConnachie A, Molyneux ME. Effect of transmission setting and mixed species infections on clinical measures of malaria in Malawi. PLoS One. 2008;3(7):e2775.

45. Black J, Hommel M, Snounou G, Pinder M. Mixed infections with Plasmodium falciparum and P malariae and fever in malaria. Lancet. 1994;343(8905):1095.

46. Betson M, Clifford S, Stanton M, Kabatereine NB, Stothard JR. Emergence of Nonfalciparum Plasmodium Infection Despite Regular Artemisinin Combination Therapy in an 18-Month Longitudinal Study of Ugandan Children and Their Mothers. J Infect Dis. 2018;217(7):1099–1109.

47. May J, Falusi AG, Mockenhaupt FP, et al. Impact of subpatent multi-species and multi-clonal plasmodial infections on anaemia in children from Nigeria. Trans R Soc Trop Med Hyg. 2000;94(4):399–403.

