## Supplemental Tables and Figures for "Epidemiology of *Plasmodium malariae* and *Plasmodium ovale* spp. in a highly malaria-endemic country: a longitudinal cohort study in Kinshasa Province, Democratic Republic of Congo"

**Short Title:** Non-falciparum malaria epidemiology in the DRC

**Supplementary Table 1. PCR primers, probes, and assay conditions**

| | | | |
|---|---|---|---|
| | **Duplex Assay: *P.* malariae-specific, and *P. ovale*-specific, 18S rRNA** | | |
| ***Adapted from:*** | Rougemont M et al. *J Clin Microbiology 2004*. 42(12):5636-43. | | |
| **Forward Primer – *Pm*** (5'->3') | CCG ACT AGG TGT TGG ATG ATA GAG TAA A | | |
| **Reverse Primer – *Pm*** (5'->3') | AAC CCA AAG ACT TTG ATT TCT CAT AA | | |
| **Forward Primer – *Po*** (5'->3') | CCG ACT AGG TTT TGG ATG AAA GAT TTT T | | |
| **Reverse Primer – *Po*** (5'->3') | AAC CCA AAG ACT TTG ATT TCT CAT AA | | |
| **Probe – *Pm*** (5'->3') | FAM-CTA TCT AAA AGA AAC ACT CAT-MGBNFQ | | |
| **Probe – *Po*** (5'->3') | VIC-CGA AAG GAA TTT TCT TAT T-MGBNFQ | | |
| **Cycling conditions:** | Temp | Duration | No. Cycles |
| | 50C | 2 min | x1 |
| | 95C | 10 min | x1 |
| | 95C | 15 sec | x45 |
| | 60C | 1 min | |
| ***Reaction conditions:*** | Roche FastStart Universal Probe Master (Rox) | | |
| | Fwd primers | 20µM | |
| | Rev primers | 20µM | |
| | Probes | 20µM | |
| | DNA | 2 µL | |
| | Total volume | 12 µL | |
| | ***P. falciparum*-specific LDH** | | |
| ***Adapted from:*** | Pickard AL et al. Antimicrob. Agents Chemo 2003. 47(8):2418-2423. | | |
| ***Forward Primer (5'->3')*** | ACGATTTGGCTGGAGCAGAT | | |
| ***Reverse Primer (5'->3')*** | TCTCTATTCCATTCTTTGTCACTCTTTC | | |
| ***Probe (5'->3')*** | FAM/ AGTAATAGTAACAGCTGGATTTACCAAGGCCCCA /TAMRA | | |
| ***Cycling conditions:*** | Temp | Duration | No. Cycles |
| | 50C | 2 min | x1 |
| | 95C | 10 min | x1 |
| | 95C | 15 sec | x40 |
| | 60C | 1 min | |
| ***Reaction conditions:*** | Roche FastStart Universal Probe Master (Rox) | | |
| | Fwd primer | 200nM | |
| | Rev primer | 200nM | |
| | Probe | 100nM | |
| | Template | 2 µl | |
| | Total volume | 12 µl | |

[1.] Duplex PCR was carried out to 45 cycle thresholds for *P. malariae* and *P. ovale* spp. detection; however, samples were considered positive if amplification under 40 Cts only, due to observed variability in this assay at later cycle thresholds.

**Supplementary Table 2. Comparison of participant characteristics between survey population and clinic subpopulation.**

| Participant Baseline Characteristics *no. (%)* | Population Type[1] | | | | |
|---|---|---|---|---|---|
| | Survey-based Population | Also in Clinic-based Subpopulation? | | | |
| | | Yes | No | p-value[3] | SMD |
| | n=1,565 participants | n=1,050 (67.1%) | n=515 (32.9%) | | |
| Age at visit (years) | | | | | |
| <5 | 302 (19.3) | 229 (21.8) | 73 (14.2) | <0.001 | 0.276 |
| 5-14 | 500 (31.9) | 354 (33.7) | 146 (28.3) | | |
| 15+ | 763 (48.8) | 467 (44.5) | 296 (57.5) | | |
| *Missing* | 0 | 0 | 0 | | |
| Sex | | | | | |
| Female | 863 (55.1) | 587 (55.9) | 276 (53.6) | 0.388 | 0.046 |
| Male | 702 (44.9) | 463 (44.1) | 239 (46.4) | | |
| *Missing* | 0 | 0 | 0 | | |
| Site | | | | | |
| Voix de Peuple | 385 (24.6) | 230 (21.9) | 155 (30.1) | <0.001 | 0.351 |
| Bu | 209 (13.4) | 171 (16.3) | 38 (7.4) | | |
| Impuru | 211 (13.5) | 136 (13.0) | 75 (14.6) | | |
| Pema | 243 (15.5) | 180 (17.1) | 63 (12.2) | | |
| Kimpoko | 167 (10.7) | 111 (10.6) | 56 (10.9) | | |
| Ngamanzo | 258 (16.5) | 167 (15.9) | 91 (17.7) | | |
| Iye | 92 (5.9) | 55 (5.2) | 37 (7.2) | | |
| *Missing* | 0 | 0 | 0 | | |
| Rurality (by health area) | | | | | |
| Urban | 385 (24.6) | 230 (21.9) | 155 (30.1) | <0.001 | 0.262 |
| Peri-urban | 517 (33.0) | 333 (31.7) | 184 (35.7) | | |
| Rural | 663 (42.4) | 487 (46.4) | 176 (34.2) | | |
| *Missing* | 0 | 0 | 0 | | |
| Wealth quintile | | | | | |
| Poorest | 318 (20.3) | 226 (21.5) | 92 (17.9) | 0.011 | 0.194 |
| Poorer | 314 (20.1) | 209 (19.9) | 105 (20.4) | | |
| Average | 311 (19.9) | 223 (21.2) | 88 (17.1) | | |
| Wealthier | 304 (19.4) | 202 (19.2) | 102 (19.8) | | |
| Wealthiest | 318 (20.3) | 190 (18.1) | 128 (24.9) | | |
| *Missing* | 0 | 0 | 0 | | |
| Fever at Baseline[2] | | | | | |
| Yes | 382 (24.5) | 272 (26.0) | 110 (21.5) | 0.052 | 0.106 |
| No | 1176 (75.5) | 774 (74.0) | 402 (78.5) | | |
| *Missing* | 7 | 4 | 3 | | |
| RDT+ at Baseline | | | | | |
| Yes | 429 (27.4) | 285 (27.2) | 144 (28.0) | 0.77 | 0.019 |
| No | 1134 (72.6) | 764 (72.8) | 370 (72.0) | | |
| *Missing* | 2 | 1 | 1 | | |
| Seasonality of visit | | | | | |
| Rainy | 1180 (75.4) | 820 (78.1) | 360 (69.9) | 0.001 | 0.188 |
| Dry | 385 (24.6) | 230 (21.9) | 155 (30.1) | | |

| Participant Baseline Characteristics *no. (%)* | Population Type[1] Survey-based Population n=1,565 participants | Also in Clinic-based Subpopulation? Yes n=1,050 (67.1%) | No n=515 (32.9%) | p-value[3] | SMD |
|---|---|---|---|---|---|
| *Missing* | 0 | 0 | 0 | | |
| Slept under bednet at Baseline | | | | | |
| Yes | 705 (45.0) | 480 (45.7) | 225 (43.7) | 0.482 | 0.041 |
| No | 860 (55.0) | 570 (54.3) | 290 (56.3) | | |
| Missing | 0 | 0 | 0 | | |
| Antimalarials taken within prior 6 months | | | | | |
| Yes | 394 (25.4) | 277 (26.6) | 117 (22.8) | 0.119 | 0.088 |
| No | 1160 (74.6) | 764 (73.4) | 396 (77.2) | | |
| Missing | 11 | 9 | 2 | | |
| *P. falciparum* PCR + | | | | | |
| Yes | 484 (30.9) | 314 (29.9) | 170 (33.0) | 0.222 | 0.067 |
| *Mixed-species* | *35 (7.2)* | *25 (8.0)* | *10 (5.9)* | *0.51* | *0.082* |
| *Single-species* | *449 (92.8)* | *289 (92.0)* | *160 (94.1)* | | |
| No | 1081 (69.1) | 736 (70.1) | 345 (67.0) | | |
| *Missing* | 0 | 0 | 0 | | |
| *P. malariae* PCR + | | | | | |
| Yes | 47 (3.0) | 33 (3.1) | 14 (2.7) | 0.753 | 0.025 |
| *Mixed-species* | *31 (66.0)* | *22 (66.7)* | *9 (64.3)* | *1* | *0.05* |
| *Single-species* | *16 (34.0)* | *11 (33.3)* | *5 (35.7)* | | |
| No | 1518 (97.0) | 1017 (96.9) | 501 (97.3) | | |
| *Missing* | 0 | 0 | 0 | | |
| *P. ovale* spp. PCR + | | | | | |
| Yes | 6 (0.4) | 5 (0.5) | 1 (0.2) | 0.67 | 0.049 |
| *Mixed-species* | *4 (66.7)* | *3 (60.0)* | *1 (100.0)* | *1* | *1.155* |
| *Single-species* | *2 (33.3)* | *2 (40.0)* | *0 (0.0)* | | |
| No | 1559 (99.6) | 1045 (99.5) | 514 (99.8) | | |
| *Missing* | 0 | 0 | 0 | | |

[1] The survey-based population comprises all participants in the study, as all participants completed the Baseline household survey. The clinic-based sub-population comprises a subset of the survey-based population who had at least 1 symptomatic clinic visit during the study period.
[2] Fever at Baseline survey was self-reported as "fever in the prior week"
[3] p-values compare baseline characteristics between those in the survey-based population who were vs. were not also included in the clinic-based symptomatic population.

**Supplemental Table 3. Baseline subject characteristics by species for the symptomatic clinic subpopulation**

| Baseline Participant Characteristics | Clinic-based Pop. | Baseline Malaria Infection by Species | | | | | |
|---|---|---|---|---|---|---|---|
| | | *P. malariae* | | *P. ovale* spp. | | *P. falciparum* | |
| | | PCR Pos. | PCR Neg. | PCR Pos. | PCR Neg. | PCR Pos. | PCR Neg. |
| no. (%) | N=1,050 | n=33 | n=1,017 | n=5 | n=1,045 | n=314 | n=736 |
| **Age (years)** | | | | | | | |
| <5 | 229 (21.8) | 4 (12.1) | 225 (22.1) | 1 (20.0) | 228 (21.8) | 49 (15.6) | 180 (24.5) |
| 5-14 | 354 (33.7) | 23 (69.7) | 331 (32.5) | 2 (40.0) | 352 (33.7) | 161 (51.3) | 193 (26.2) |
| 15+ | 467 (44.5) | 6 (18.2) | 461 (45.3) | 2 (40.0) | 465 (44.5) | 104 (33.1) | 363 (49.3) |
| **Sex** | | | | | | | |
| Female | 587 (55.9) | 19 (57.6) | 568 (55.9) | 5 (100.0) | 582 (55.7) | 165 (52.5) | 422 (57.3) |
| Male | 463 (44.1) | 14 (42.4) | 449 (44.1) | 0 (0.0) | 463 (44.3) | 149 (47.5) | 314 (42.7) |
| **Urbanicity** | | | | | | | |
| Rural | 487 (46.4) | 21 (63.6) | 466 (45.8) | 5 (100.0) | 482 (46.1) | 197 (62.7) | 290 (39.4) |
| Peri-urban | 333 (31.7) | 10 (30.3) | 323 (31.8) | 0 (0.0) | 333 (31.9) | 110 (35.0) | 223 (30.3) |
| Urban | 230 (21.9) | 2 (6.1) | 228 (22.4) | 0 (0.0) | 230 (22.0) | 7 (2.2) | 223 (30.3) |
| **Fever (≤1 week)** | | | | | | | |
| Yes | 272 (26.0) | 9 (28.1) | 263 (25.9) | 4 (80.0) | 268 (25.7) | 110 (35.3) | 162 (22.1) |
| **RDT+** | | | | | | | |
| Yes | 285 (27.2) | 20 (62.5) | 265 (26.1) | 2 (40.0) | 283 (27.1) | 235 (75.1) | 50 (6.8) |
| **Bed Net Use (Prior Night)** | | | | | | | |
| Yes | 480 (45.7) | 9 (27.3) | 471 (46.3) | 2 (40.0) | 478 (45.7) | 127 (40.4) | 353 (48.0) |
| **Symptoms in Prior 6 mon.** | | | | | | | |
| Yes | 269 (25.7) | 7 (21.9) | 262 (25.8) | 3 (60.0) | 266 (25.5) | 81 (26.0) | 188 (25.6) |
| **Tx with Antimalarials in Prior 6 mon.** | | | | | | | |
| Yes | 277 (26.6) | 7 (21.9) | 270 (26.8) | 1 (20.0) | 276 (26.6) | 76 (24.4) | 201 (27.6) |
| **Wealth Category** | | | | | | | |
| Poorest | 226 (21.5) | 5 (15.2) | 221 (21.7) | 1 (20.0) | 225 (21.5) | 93 (29.6) | 133 (18.1) |
| Poorer | 209 (19.9) | 14 (42.4) | 195 (19.2) | 0 (0.0) | 209 (20.0) | 78 (24.8) | 131 (17.8) |
| Average | 223 (21.2) | 6 (18.2) | 217 (21.3) | 2 (40.0) | 221 (21.1) | 69 (22.0) | 154 (20.9) |
| Wealthier | 202 (19.2) | 6 (18.2) | 196 (19.3) | 2 (40.0) | 200 (19.1) | 69 (22.0) | 133 (18.1) |
| Wealthiest | 190 (18.1) | 2 (6.1) | 188 (18.5) | 0 (0.0) | 190 (18.2) | 5 (1.6) | 185 (25.1) |

**Supplemental Table 4. Participant characteristics across follow-up visits (Survey population)**

| Participant characteristics at each visit *no. (%)* | Household Survey Visits (*Active surveillance*) Baseline n=1,565 | Follow-up 1 n=1,447 | Follow-up 2 n=1,367 | Follow-up 3 n=1,303 | p-value[3] |
|---|---|---|---|---|---|
| No. subjects | 1,565 | 1,447 | 1,367 | 1,303 | |
| Age at visit (years) | | | | | |
| <5 | 302 (19.3) | 271 (18.7) | 225 (16.5) | 185 (14.2) | 0.009 |
| 5-14 | 500 (31.9) | 480 (33.2) | 443 (32.4) | 453 (34.8) | |
| 15+ | 763 (48.8) | 696 (48.1) | 699 (51.1) | 665 (51.0) | |
| *Missing* | *0* | *0* | *0* | *0* | |
| Sex | | | | | |
| Female | 863 (55.1) | 800 (55.3) | 757 (55.4) | 707 (54.3) | 0.935 |
| Male | 702 (44.9) | 647 (44.7) | 610 (44.6) | 596 (45.7) | |
| *Missing* | *0* | *0* | *0* | *0* | |
| Rurality (by health area) | | | | | |
| Urban | 385 (24.6) | 342 (23.6) | 307 (22.5) | 300 (23.0) | 0.384 |
| Peri-urban | 517 (33.0) | 487 (33.7) | 457 (33.4) | 402 (30.9) | |
| Rural | 663 (42.4) | 618 (42.7) | 603 (44.1) | 601 (46.1) | |
| *Missing* | *0* | *0* | *0* | *0* | |
| Wealth quintile | | | | | |
| Poorest | 318 (20.3) | 294 (20.3) | 290 (21.2) | 274 (21.0) | 0.994 |
| Poorer | 314 (20.1) | 288 (19.9) | 277 (20.3) | 264 (20.3) | |
| Average | 311 (19.9) | 301 (20.8) | 292 (21.4) | 271 (20.8) | |
| Wealthier | 304 (19.4) | 284 (19.6) | 255 (18.7) | 249 (19.1) | |
| Wealthiest | 318 (20.3) | 280 (19.4) | 253 (18.5) | 245 (18.8) | |
| *Missing* | *0* | *0* | *0* | *0* | |
| Fever[1] | | | | | |
| Yes | 382 (24.5) | 173 (12.0) | 249 (18.2) | 120 (9.2) | <0.001 |
| No | 1176 (75.5) | 1273 (88.0) | 1118 (81.8) | 1183 (90.8) | |
| *Missing* | *7* | *1* | *0* | *0* | |
| RDT+ | | | | | |
| Yes | 429 (27.4) | 396 (27.4) | 511 (37.4) | 307 (23.6) | <0.001 |
| No | 1134 (72.6) | 1051 (72.6) | 856 (62.6) | 995 (76.4) | |
| *Missing* | *2* | *0* | *0* | *1* | |
| Slept under bednet the prior night | | | | | |
| Yes | 705 (45.0) | 810 (56.0) | 732 (53.5) | 589 (45.2) | <0.001 |
| No | 860 (55.0) | 637 (44.0) | 635 (46.5) | 714 (54.8) | |
| *Missing* | *0* | *0* | *0* | *0* | |
| Seasonality of visit | | | | | |
| Rainy | 1180 (75.4) | 0 (0.0) | 1365 (99.9) | 19 (1.5) | <0.001 |
| Dry | 385 (24.6) | 1447 (100.0) | 2 (0.1) | 1284 (98.5) | |
| *Missing* | *0* | *0* | *0* | *0* | |
| Antimalarials taken within prior 6 months[2] | | | | | |
| Yes | 394 (25.4) | 559 (38.9) | 625 (46.1) | 569 (46.6) | <0.001 |
| No | 1160 (74.6) | 877 (61.1) | 732 (53.9) | 652 (53.4) | |
| *Missing* | *11* | *11* | *10* | *82* | |
| *P. falciparum* PCR + | | | | | |
| Yes | 484 (30.9) | 512 (35.5) | 538 (39.6) | 442 (34.1) | <0.001 |

| Participant characteristics at each visit *no. (%)* | Household Survey Visits (*Active surveillance)* Baseline n=1,565 | Follow-up 1 n=1,447 | Follow-up 2 n=1,367 | Follow-up 3 n=1,303 | p-value[3] |
|---|---|---|---|---|---|
| *Mixed-species* | *35 (7.2)* | *42 (8.2)* | *64 (11.9)* | *47 (10.6)* | *0.043* [4] |
| *Single-species* | *449 (92.8)* | *470 (91.8)* | *474 (88.1)* | *395 (89.4)* | |
| No | 1081 (69.1) | 930 (64.5) | 820 (60.4) | 853 (65.9) | |
| *Missing* | *0* | *5* | *9* | *8* | |
| *P. malariae* PCR + | | | | | |
| Yes | 47 (3.0) | 35 (2.4) | 56 (4.1) | 48 (3.7) | 0.055 [4] |
| *Mixed-species* | *31 (66.0)* | *24 (68.6)* | *44 (78.6)* | *38 (79.2)* | *0.346* [4] |
| *Single-species* | *16 (34.0)* | *11 (31.4)* | *12 (21.4)* | *10 (20.8)* | |
| No | 1518 (97.0) | 1408 (97.6) | 1300 (95.9) | 1247 (96.3) | |
| *Missing* | *0* | *4* | *11* | *8* | |
| *P. ovale* spp. PCR + | | | | | |
| Yes | 6 (0.4) | 27 (1.9) | 27 (2.0) | 18 (1.4) | <0.001 [4] |
| *Mixed-species* | *4 (66.7)* | *20 (74.1)* | *24 (88.9)* | *13 (72.2)* | *0.358* [4] |
| *Single-species* | *2 (33.3)* | *7 (25.9)* | *3 (11.1)* | *5 (27.8)* | |
| No | 1559 (99.6) | 1416 (98.1) | 1329 (98.0) | 1277 (98.6) | |
| *Missing* | *0* | *4* | *11* | *8* | |

[1] Fever was self-reported as "fever in the prior week"

[2] Self-reported use of antimalarials in prior 6 months.

[3] Categorical variables were statistically compared using chi-squared tests; continuous variables were compared using the Kruskal-Wallis test of medians to account for non-normality. Missing data were excluded from statistical tests.

[4] p-values tested using Fisher's exact testing due to small cell sizes.

**Supplemental Table 5. Participant characteristics across follow-up (Clinic Subpopulation)**

| *Participant* characteristics at visits<br>*no. (%)* | Clinic Visits<br>Time from Baseline Visit[2]<br>All visits within first 12 months<br>n=1297 | All visits between 12 to 24 months<br>n=1203 | All visits after 24 months<br>n=907 | p-value[3] |
|---|---|---|---|---|
| No. subjects with visits | 732 | 648 | 510 | |
| Age at visit (years) | | | | |
| <5 | 353 (27.2) | 305 (25.4) | 172 (19.0) | <0.001 |
| 5-14 | 481 (37.1) | 434 (36.1) | 386 (42.6) | |
| 15+ | 463 (35.7) | 462 (38.5) | 348 (38.4) | |
| *Missing* | *0* | *2* | *1* | |
| Sex | | | | |
| Female | 741 (57.1) | 726 (60.3) | 514 (56.7) | 0.153 |
| Male | 556 (42.9) | 477 (39.7) | 393 (43.3) | |
| *Missing* | *0* | *0* | *0* | |
| Site | | | | |
| Voix de Peuple | 182 (14.0) | 213 (17.7) | 109 (12.0) | <0.001 |
| Bu | 232 (17.9) | 305 (25.4) | 236 (26.0) | |
| Impuru | 177 (13.6) | 139 (11.6) | 73 (8.0) | |
| Pema | 261 (20.1) | 203 (16.9) | 162 (17.9) | |
| Kimpoko | 217 (16.7) | 114 (9.5) | 104 (11.5) | |
| Ngamanzo | 181 (14.0) | 187 (15.5) | 190 (20.9) | |
| Iye | 47 (3.6) | 42 (3.5) | 33 (3.6) | |
| *Missing* | *0* | *0* | *0* | |
| Rurality (by health area) | | | | |
| Urban | 182 (14.0) | 213 (17.7) | 109 (12.0) | <0.001 |
| Peri-urban | 445 (34.3) | 343 (28.5) | 327 (36.1) | |
| Rural | 670 (51.7) | 647 (53.8) | 471 (51.9) | |
| *Missing* | *0* | *0* | *0* | |
| Wealth quintile | | | | |
| Poorest | 288 (22.2) | 235 (19.5) | 201 (22.2) | <0.001 |
| Poorer | 267 (20.6) | 309 (25.7) | 230 (25.4) | |
| Average | 332 (25.6) | 279 (23.2) | 232 (25.6) | |
| Wealthier | 267 (20.6) | 192 (16.0) | 156 (17.2) | |
| Wealthiest | 143 (11.0) | 188 (15.6) | 88 (9.7) | |
| *Missing* | *0* | *0* | *0* | |
| Fever[1] | | | | |
| Yes | 649 (75.1) | 384 (48.7) | 274 (44.4) | <0.001 [4] |
| No | 215 (24.9) | 404 (51.3) | 343 (55.6) | |
| *Missing* | *433* | *415* | *290* | |
| RDT+ | | | | |
| Yes | 1145 (88.4) | 948 (79.0) | 687 (76.1) | <0.001 [4] |
| No | 150 (11.6) | 252 (21.0) | 216 (23.9) | |
| *Missing* | *2* | *3* | *4* | |
| Anemia | | | | |
| Severe | 48 (4.1) | 32 (2.7) | 17 (1.9) | 0.009 |
| Moderate | 249 (21.4) | 230 (19.4) | 167 (18.8) | |
| Mild | 202 (17.4) | 242 (20.4) | 197 (22.2) | |
| Not anemic | 662 (57.0) | 681 (57.5) | 506 (57.0) | |

| *Participant* characteristics at visits *no. (%)* | Clinic Visits<br>Time from Baseline Visit[2]<br>**All visits within first 12 months**<br>n=1297 | **All visits between 12 to 24 months**<br>n=1203 | **All visits after 24 months**<br>n=907 | p-value[3] |
|---|---|---|---|---|
| *Missing* | *136* | *18* | *20* | |
| Seasonality of visit | | | | |
| Rainy | 885 (68.2) | 758 (63.0) | 411 (45.3) | <0.001 [4] |
| Dry | 412 (31.8) | 445 (37.0) | 496 (54.7) | |
| *Missing* | *0* | *0* | *0* | |
| *P. falciparum* PCR + | | | | |
| Yes | 829 (64.6) | 636 (53.5) | 544 (62.0) | <0.001 |
| *Mixed-species* | *53 (6.4)* | *42 (6.6)* | *41 (7.5)* | *0.694* [4] |
| *Single-species* | *775 (93.6)* | *593 (93.4)* | *503 (92.5)* | |
| No | 455 (35.4) | 552 (46.5) | 334 (38.0) | |
| *Missing* | *13* | *15* | *29* | |
| *P. malariae* PCR + | | | | |
| Yes | 53 (4.1) | 44 (3.7) | 38 (4.3) | 0.757 [4] |
| *Mixed-species* | *35 (66.0)* | *25 (56.8)* | *29 (76.3)* | *0.187* [4] |
| *Single-species* | *18 (34.0)* | *19 (43.2)* | *9 (23.7)* | |
| No | 1230 (95.9) | 1142 (96.3) | 842 (95.7) | |
| *Missing* | *14* | *17* | *27* | |
| *P. ovale* spp. PCR + | | | | |
| Yes | 34 (2.7) | 41 (3.5) | 20 (2.3) | 0.255 [4] |
| *Mixed-species* | *19 (55.9)* | *21 (51.2)* | *12 (60.0)* | *0.864* [4] |
| *Single-species* | *15 (44.1)* | *20 (48.8)* | *8 (40.0)* | |
| No | 1249 (97.3) | 1145 (96.5) | 860 (97.7) | |
| *Missing* | *14* | *17* | *27* | |

[1] Fever was measured at time of clinic visit.
[2] Clinic visits could continue past the end of active surveillance till the end of 2017, for a total of 34 months from the first baseline visit. Clinic visits after 24 months occurred after all household surveys had concluded.
[3] Categorical variables were statistically compared using chi-squared tests; continuous variables were compared using the Kruskal-Wallis test of medians to account for non-normality. Missing data were excluded from statistical tests.
[4] p-values tested using Fisher's exact testing due to small cell sizes.

**Supplemental Figure 1 A-B. ) Factors associated with *P. malariae* and *P. ovale* spp. infection prevalence, compared to *P. falciparum,* stratified by study population**.

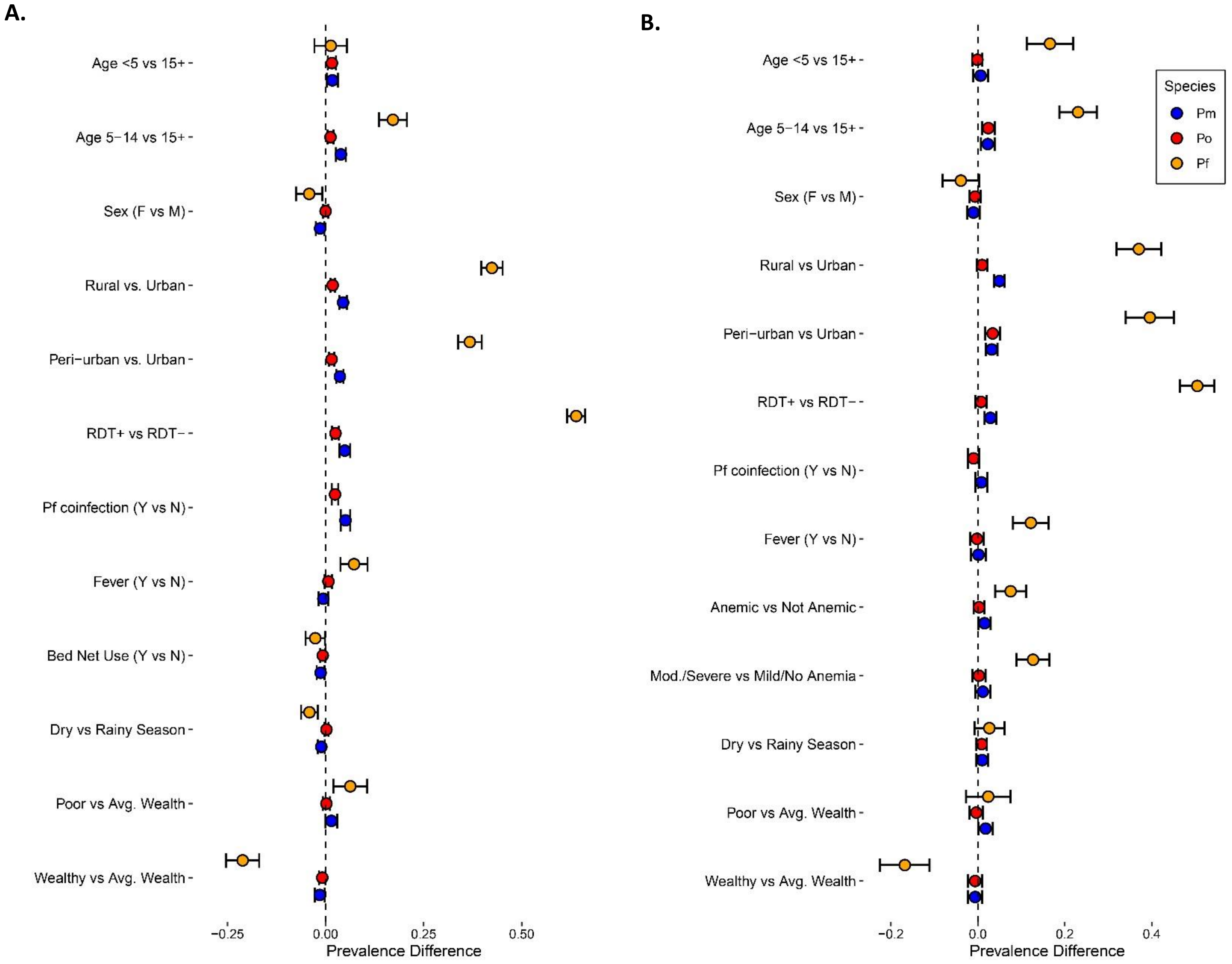


A) Factors associated with non-falciparum infection prevalence compared to *P. falciparum* infection, at survey visits (baseline and three follow-up surveys). B) Factors associated with non-falciparum infection prevalence compared to *P. falciparum* infection, at clinic visits throughout follow-up.

**Supplemental Figures 2 A-D.** ***P. malariae*** **and** ***P. ovale*** **spp. infections throughout the study period, within the Total Population, encompassing infections detected at all touch points in the study (survey + clinic visits).**

**A.**

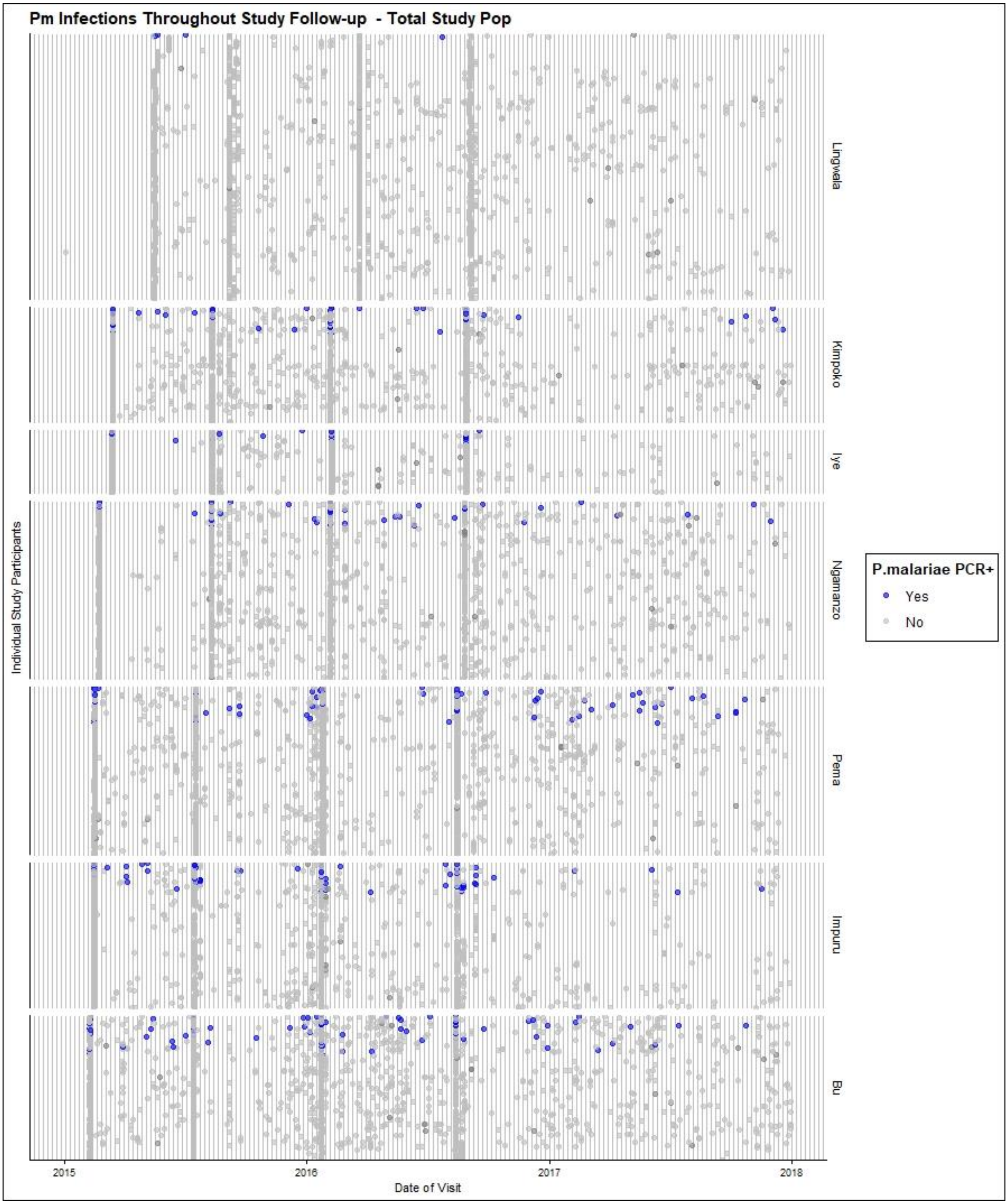


A) *P. malariae* infections detected at study visits throughout follow-up, among all subjects in the Total Population. Rows represent individual subjects, sorted by frequency of PCR+ *P. malariae* infections throughout the full study period.

**B**

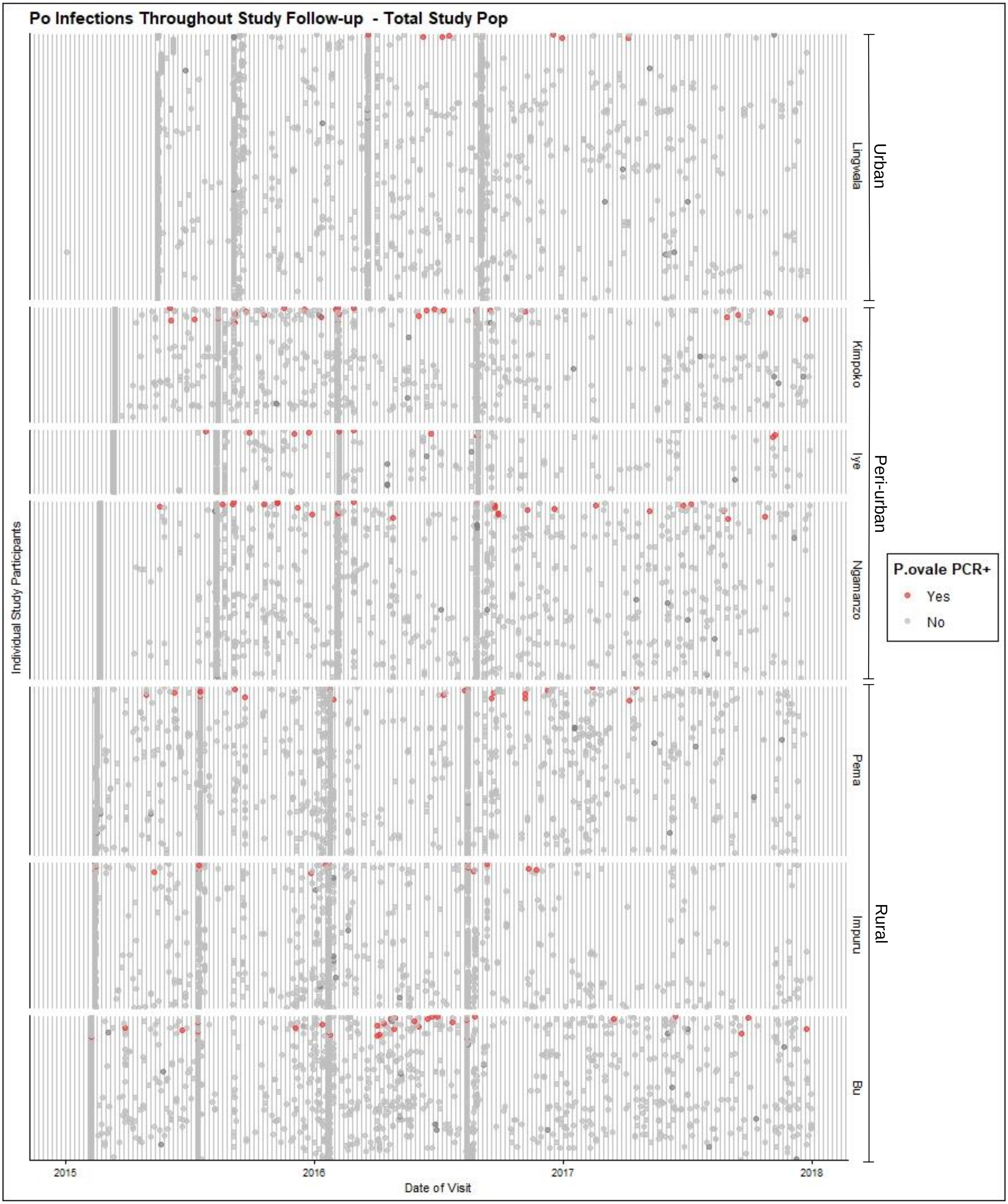


B) *P. ovale* spp. infections detected at study visits throughout follow-up, among all subjects in the Total Population. Rows represent individual subjects, sorted by frequency of PCR+ *P. ovale* spp. infections throughout the full study period.

C

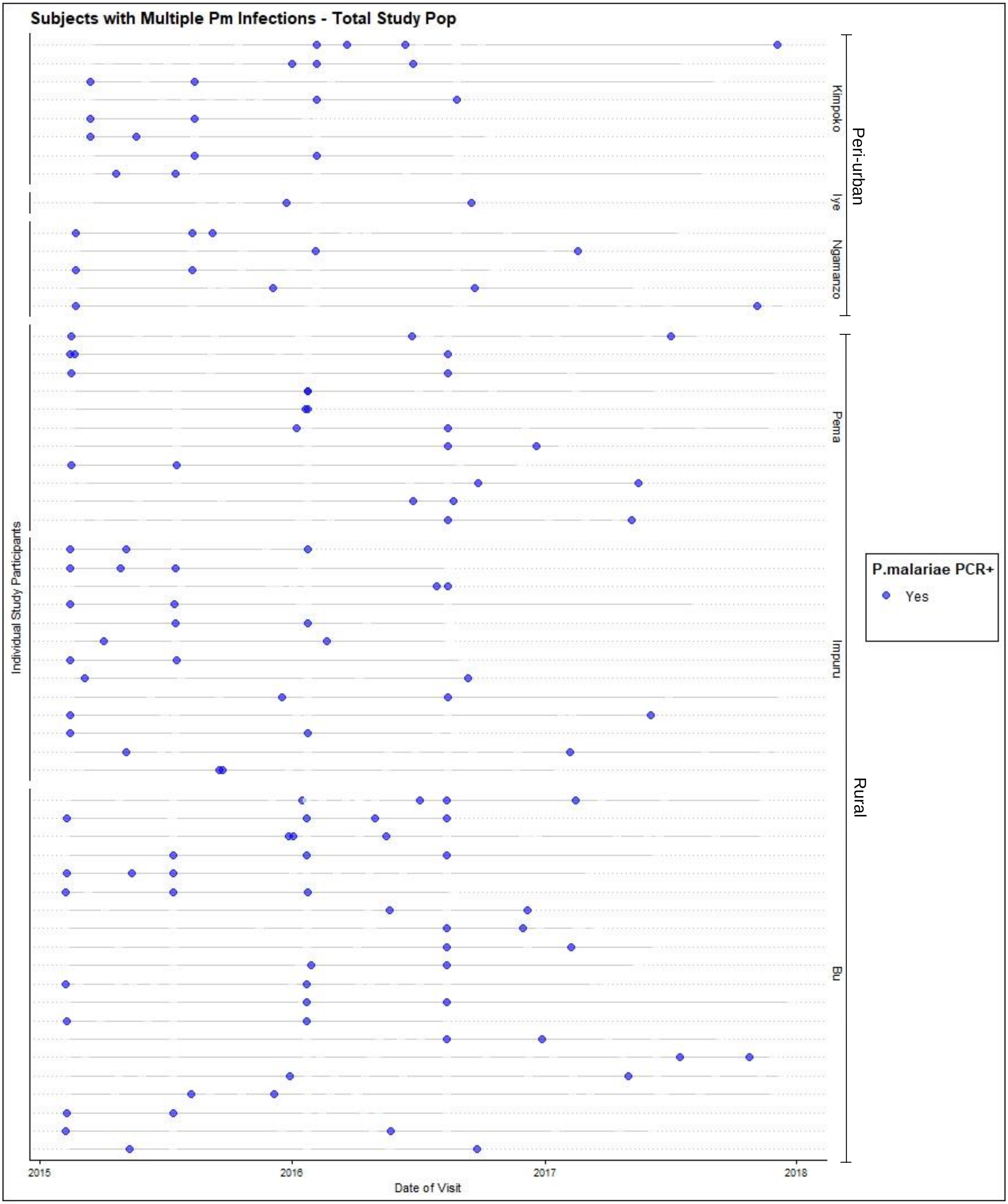


C) Multiple *P. malariae* infections detected at study visits throughout follow-up, among all subjects in the Total Population who had at least one *P.* malariae infection during the study. Rows represent individual subjects, sorted by frequency of PCR+ *P. malariae* spp. infections throughout the full study period.

**D**

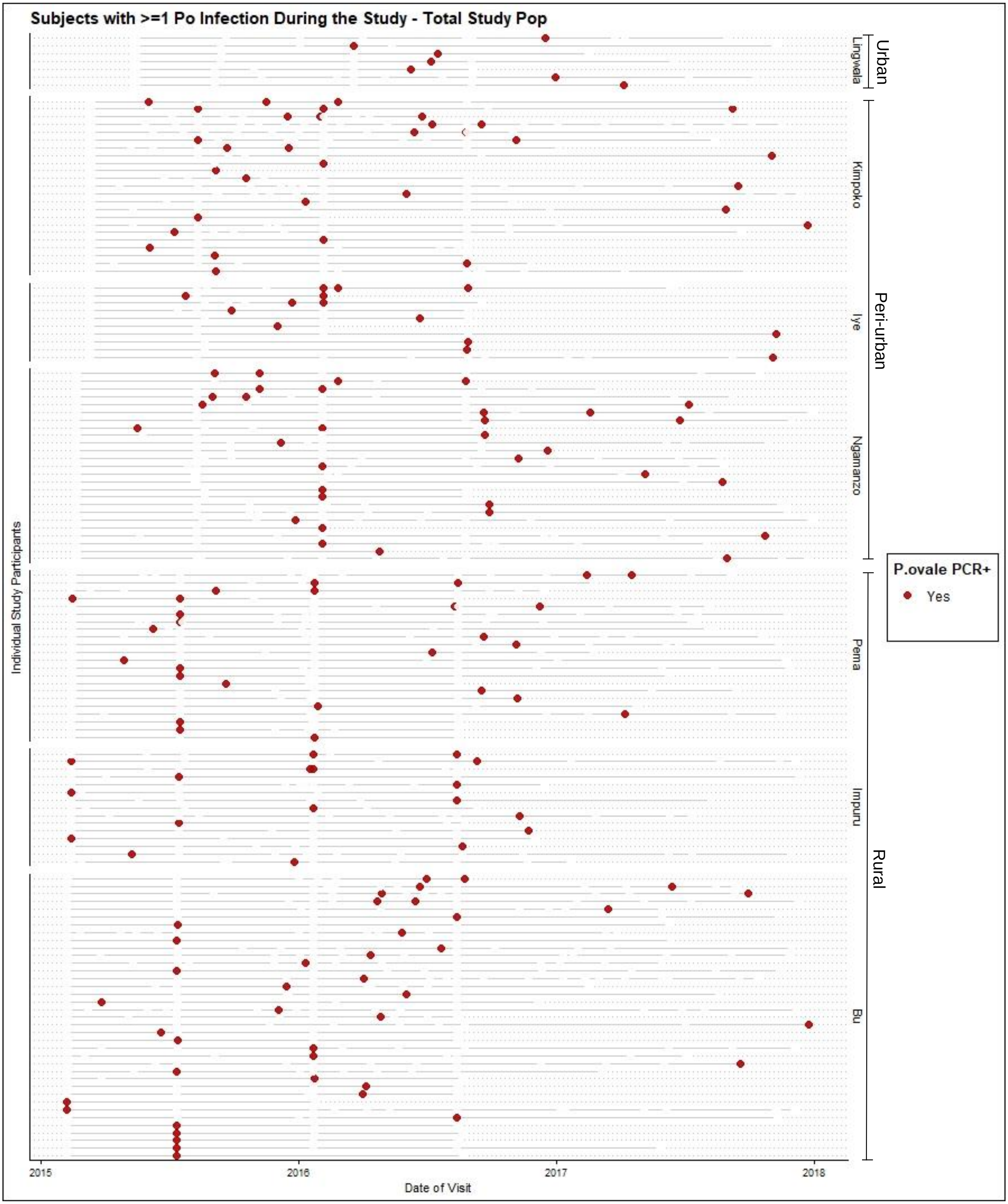


D) Multiple *P. ovale* spp. infections detected at study visits throughout follow-up, among all subjects in the Total Population <u>who had at least one *P.* ovale spp. infection during the study</u>. Rows represent individual subjects, sorted by frequency of PCR+ *P. ovale* spp. infections throughout the full study period.

**Supplemental Table 6. Estimated parasitemias (p/µL) by malaria species**

| Parasitemia Estimates (p/uL) | | **Survey Population** – *Household Survey Visits* | | | | | |
|---|---|---|---|---|---|---|---|
| | | n (infect.) | median | IQR | min - max | p-value | n excluded (rehydrated)‡ |
| **Pm (p/uL)** | Total | 175 | 22.4 | 8.5-72.2 | 0.8-358,246 | | 11 |
| | Mixed | 129 | 21.1 | 8.5-60.1 | 1.4-358,246 | 0.259 | 8 |
| | Mono | 46 | 28.8 | 9.2-125 | 0.8-688 | | 3 |
| **Po (p/uL)** | Total | 71 | 5.8 | 2.0-28.0 | 0.6-106,476 | | 7 |
| | Mixed | 56 | 5.7 | 2.3-23.0 | 0.6-106,476 | 0.662 | 5 |
| | Mono | 15 | 9.5 | 1.7-92.2 | 0.8-252 | | 2 |
| **Pf (p/uL)** | Total | 1760 | 52.6 | 8.2-343.3 | 0.6-268250.0 | | 216 |
| | Mixed | 176 | 120.5 | 31.9-496.3 | 0.8-22925.0 | <0.001 | 12 |
| | Mono | 1584 | 46.9 | 7.4-311.1 | 0.6-268250.0 | | 204 |

| Parasitemia Estimates (p/uL) | | **Clinic sub-Population** – *Symptomatic Clinic Visits* | | | | | |
|---|---|---|---|---|---|---|---|
| | | n (infect.) | median | IQR | min - max | p-value | n excluded (rehydrated) |
| **Pm (p/uL)** | Total | 132 | 36.5 | 4.7-182 | 0.6-23,288 | | 3 |
| | Mixed | 88 | 31.8 | 3.6-157 | 0.6-2,432 | 0.239 | 1 |
| | Mono | 44 | 51.5 | 15.2-204 | 1.3-23,288 | | 2 |
| **Po (p/uL)** | Total | 93 | 17.7 | 4.6-65.8 | 0.2-2,875 | | 2 |
| | Mixed | 51 | 17.8 | 4.9-68.6 | 0.3-1,447 | 0.740 | 1 |
| | Mono | 42 | 16.7 | 4.5-65.5 | 0.2-2,875 | | 1 |
| **Pf (p/uL)** | Total | 1970 | 2644.3 | 113.2-16931.1 | 0.6-1,165,100 | | 39 |
| | Mixed | 134 | 508.7 | 58.8-8387.1 | 2.1-119,100 | 0.001 | 2 |
| | Mono | 1834 | 2897 | 126.5-18058.3 | 0.6-1,165,100 | | 37 |

| Parasitemia Estimates (p/uL) | | **Total Population = Survey and Clinic Visits** | | | | | |
|---|---|---|---|---|---|---|---|
| | | n (infect.) | median | IQR | min - max | p-value | n excluded (rehydrated) |
| **Pm (p/uL)** | Total | 307 | 25.7 | 7.7-119 | 0.6-358,246 | | 14 |
| | Mixed | 217 | 22.4 | 6.9-108 | 0.6-358,246 | 0.071 | 9 |
| | Mono | 90 | 36.5 | 11.8 -187 | 0.8-23,288 | | 5 |
| **Po (p/uL)** | Total | 164 | 10.2 | 2.7-47.4 | 0.2-106,476 | | 9 |
| | Mixed | 107 | 10.8 | 2.8-36.4 | 0.3-106.476 | 0.465 | 6 |
| | Mono | 57 | 15.8 | 2.2-65.8 | 0.2-2,875 | | 3 |
| **Pf (p/uL)** | Total | 3730 | 266.5 | 18.8-4525.5 | 0.6-1,165,100 | | 255 |
| | Mixed | 310 | 190 | 40.7-1427.4 | 0.8-119,100 | 0.209 | 14 |
| | Mono | 3418 | 279.6 | 17.4-5014.6 | 0.6-1,165,100 | | 241 |
